# Quantifiable identification of flow-limited ventilator dyssynchrony with the deformed lung ventilator model

**DOI:** 10.1101/2023.06.16.23291492

**Authors:** Deepak. K. Agrawal, Bradford J. Smith, Peter D. Sottile, George Hripcsak, David J. Albers

## Abstract

**Objective:** Ventilator dyssynchrony (VD) is potentially harmful to patients with or at risk for acute respiratory distress syndrome (ARDS). In addition to injury solely caused by the ventilator, ventilator-induced lung injury may be instigated and exacerbated by patient respiratory efforts. Automated detection of VD from ventilator waveforms is challenging, and efforts have been made on a human-guided ML algorithm to detect some types of VD. We currently lack a methodological ability to define sub-breath phenotypes of VD that quantify severity anchored to physiologic understanding that could be used to relate VD to damage and guide ventilator management.

**Materials and Methods:** A mathematical model is developed that represents the pressure and volume waveform signals of a breath into several pathophysiological temporal features observed in ventilator waveforms and then deformation terms are added corresponding to hypothesized flow-limited (FL) dyssynchronous breaths. Model parameters are estimated at the resolution of a single breath using a deterministic, multivariate, constrained interior-point method to create a parametric representation of breaths. Differential estimates of different FL-VD breaths are used to create severity metrics for FL-VD breaths and their associations with the ventilator settings and healthcare interventions are analyzed.

**Results:** A total of 93,007 breaths were analyzed from the raw ventilator waveform dataset of 13 intensive care unit patients who met inclusion criteria. A quantitative method was developed to determine the continuously varying FL-VD severity for each breath and was successfully applied to a cohort of patient-ventilator waveform data. Additionally, cross-validation, using a previously developed ML categorical VD identification algorithm, produced an area under the receiver operator curve of 0.97.

**Discussion & Conclusion:** The VD-deformed lung ventilator (VD-DLV) model accurately detects FL-VD breaths and is able to quantify the severity of patient effort during patient-ventilator interaction. The presence and severity of deviations from normal are modeled in a way that is based on physiological hypotheses of lung damage and ventilator interactions. Therefore, the computed phenotypes have the predictive power to determine how the healthcare variables are associated with FL-VD breaths. This work paves the way for a large-scale study of VD causes and effects by identifying and quantifying VD breaths using the VD-DLV model.

## INTRODUCTION

Mechanical ventilation (MV) is one of the most important life-saving interventions for those unable to achieve the gas exchange necessary to maintain life. However, MV can cause ventilator-induced lung injury (VILI), especially in patients with or at risk for acute respiratory distress syndrome (ARDS) [1-3]. A synchronized patient-ventilator interaction, where patient inspiratory efforts trigger appropriately-timed ventilator support, is necessary for optimal performance of the MV while minimizing VILI [4, 5]. However, achieving patient-ventilator synchrony is often challenging to achieve due to the complex interplay between ventilator mechanics, rapidly changing underlying pulmonary physiology and healthcare interventions.

One pathway for VILI leading to poor outcomes occurs when patient respiratory efforts and ventilator support are miss-timed - a process termed ventilator dyssynchrony (VD) [6-10]. Dyssynchrony is a heterogeneous phenomenon with different phenotypes defined by the timing of the respiratory effort with respect to ventilator support. The presence of VD is associated with high mortality in ARDS, and there are hypothesized pathways for damage with different types of VD [7]. ARDS itself is common and deadly: excluding COVID-19, 35-45% of the 10% of ICU patients with ARDS, or 3.5-4.5% of all ICU patients, will die with ARDS. The true impact of ARDS on mortality is likely greater because while few patients with ARDS die from refractory hypoxemia, ARDS has a profoundly negative impact on other organ systems [11]. Decoupling VD, VILI and ARDS is difficult because they co-occur and are hypothesized to cause and exacerbate each other. While VD is thought to significantly worsen ARDS and VILI, the exact mechanisms and the types of VD that cause the worst injury are not well understood [5]. Similarly, quantifying the relationship between VD and VILI is challenging because of inter-breath heterogeneity and the inability to define lung state by breath. In short, a significant roadblock to advancing ventilator management are the missing VD breath phenotype labels or definitions using variables that relate to physiological and ventilator setting features that can lead to new, more protective interventions. A quantitative link that can inform how VD drives VILI and mortality would significantly advance MV management and patient care.

In clinical settings, the primary way to detect VD is by manually examining ventilator waveforms such as pressure-time, volume-time, and flow-time signals at the bedside [12-15]. This process is labor-intensive and full of diagnostic inaccuracies because the recognition ability of the individual clinician drives the outcome [16]. Additionally, in the manual process, other relevant information is often missed, such as the relation between varying types and frequencies of VD breaths over time, ventilator settings, and other time-sensitive healthcare variables [17-19]. Some of these limitations are being addressed using rule-based machine learning (ML) algorithms to identify different VD types [20-24]. These approaches require manually selecting important waveform features from noisy and heterogeneous signals, and hence accurately extracting features from respiratory sequences is challenging. Due to this, current ML-based algorithms only categorically detect some forms of VD and accuracy may be limited [5]. In addition, these ML approaches do not specify the severity of VD, which exists on a continuum. This is important because different magnitudes of effort may lead to differential effects on the lung with, for example, vigorous inspirations leading to VILI while less severe efforts may not damage the lungs. This severity stratification is critical to determining the association between VD and lung damage. Furthermore, these approaches do not reveal the waveform characteristics used to classify the VD type, and those data may serve as markers of damage pathways (e.g., overdistension) and correlate with outcomes.

Physiological mathematical models are an alternative to ML but can struggle with knowledge extraction from the clinical data as they include only half of the system, the lung, and do not model the ventilator or the human-ventilator interactions. Because of this, they can miss clinically meaningful dynamics such as respiratory efforts and patient-ventilator interactions. For example, in compartment-based models, it remains unknown which important clinically observable features of lung damage can be represented [25, 26]. While other mathematical models can capture the lung damage from the ventilator waveform data as long as patient-ventilator interactions are synchronous [27, 28]. However, in clinical settings, the patient often interacts with the ventilator in a dyssynchronous manner, which ultimately drives VD [5]. Due to these challenges, in existing clinical practice ventilator waveforms are often underutilized to troubleshoot and optimize patient–ventilator interactions and extract information about pulmonary physiology.

To understand the pathophysiological relationship between VD, VILI, and ARDS, a quantitative method is needed to demarcate VD by type and severity to link those characteristics to healthcare interventions, lung dysfunction and patient outcomes. This will allow determination of the circumstances leading to the worst damage and outcomes. For example, some types of VD may do minor damage when the lungs are healthy but be injurious when the lungs are already compromised [17, 29]. In contrast, other kinds of VD may always cause damage or could even be lung protective. In addition, some types of initial lung injury might be more susceptible to particular types of VD than others. Developing this understanding requires a systematic framework to define quantifiable phenotypes of dyssynchrony at the resolution of a single breath, anchored to physiologic understanding that could be used to guide ventilator management (Fig. 1a). In such a framework, VD can be conceptualized as deviation from ‘ideal’ pressure and volume waveforms. This is out of reach with current fully mechanistic models, and while ML approaches are useful, they are not hypothesis-driven and hence lack explainability that can lead to new interventions.

**Figure 1:**
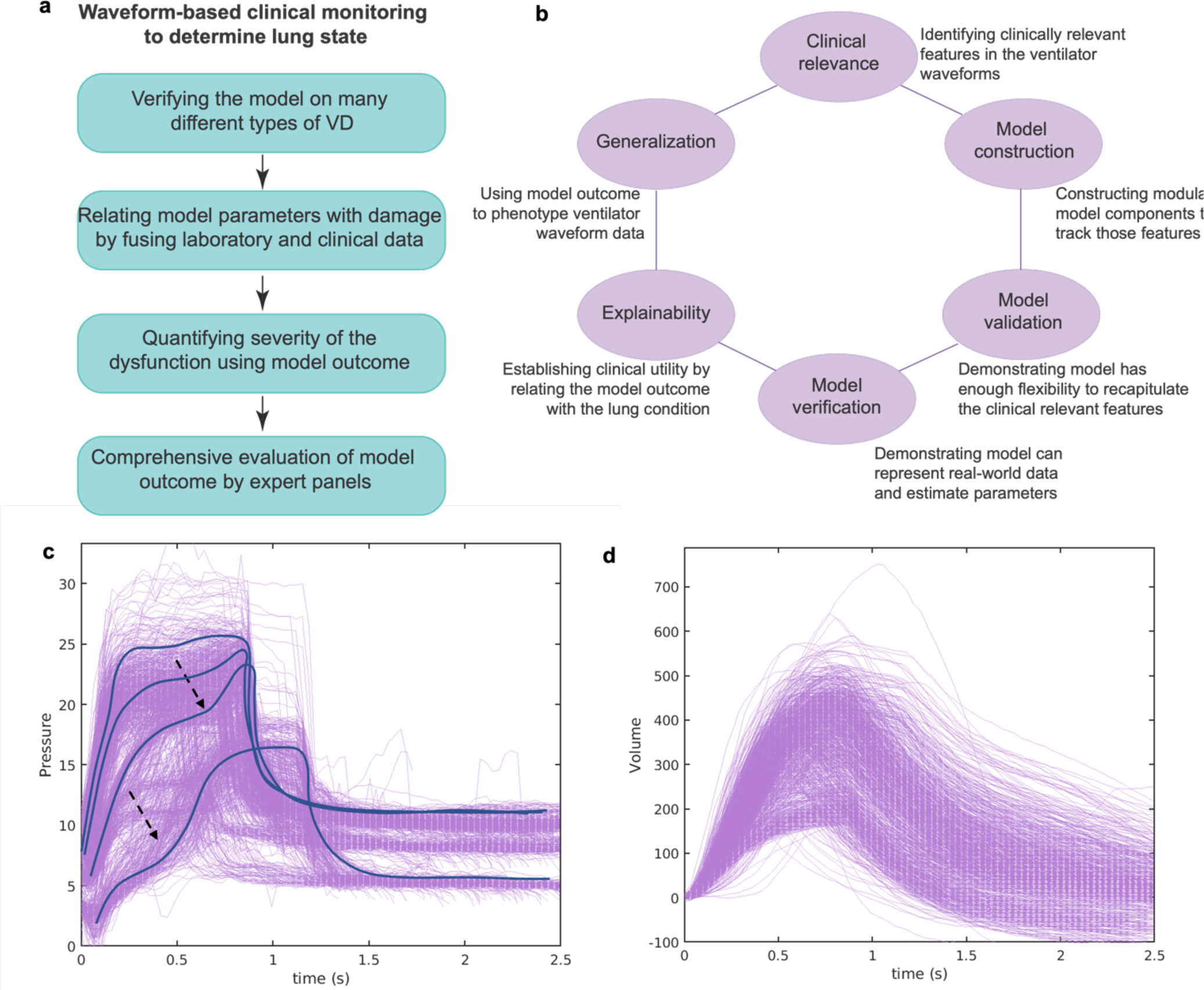
**(a)** Describing a waveform-based clinical monitoring framework for identifying and quantifying different types of VD breaths. **(b)** A schematic diagram summarizing the earlier work where we developed a physiologically anchored data-driven model, intending to reproduce ventilator waveform data and, in doing so, infer injured lung dynamics [30]. The model takes, as a baseline, ideal pressure and volume waveforms and then adds functional terms that correspond to particular deformities in the waveform, which are often linked to the physiological states of the lung by clinicians [19]. **(c-d)** Representative recorded **(c)** pressure and **(d)** volume waveform signals of normal (NL) and flow-limited (FL) breaths for an ICU patient ID 104 (see Methods). The scooping (black arrow) in the pressure signal coincides with patient effort when the flow is not adequate and could be used to identify and quantify the FL-VD breaths [32]. During the treatment, the ventilator was operated in Adaptive Pressure Ventilation - Controlled Mechanical Ventilation (APVCMV) mode [16].

A solution to this problem may lie in a hybrid approach where the ventilator mechanics, the lungs, and their coupled interaction are integrated into a unified modeling framework. This approach lives between the fully mechanistic lung modeling approach, where the observed physiology must emerge from the proposed lung mechanics, and a machine learning approach, where the model is flexible enough to estimate every feature and must discern which features are important. Using this approach, we have already developed a physiologically linked and data-driven lung ventilator model where model parameters control features of the pressure and volume waveform signals that are hypothesized to be related to specific physiology and pathophysiology [30]. We refer to that model framework as the damaged informed lung ventilator (DILV) model. Our prior work showed that the model can differentiate between lung injury states using a single breath from a mouse or human, and the model accuracy compares favorably to the single-compartment model (Fig. 1b). We did not, however, accommodate patient-ventilator interactions in that model [30].

In the current study, we take the first steps toward constructing a new, modular modeling framework that can identify specific features in the pressure waveform that arises when the ventilator does not deliver support (flow) proportional to the patient’s demand. This is often described as flow-limited ventilator dyssynchrony (FL-VD) and is characterized by a “scooping” of airway pressure (Fig. 1c, black arrow). At the same time, the shape of the volume signal is relatively unchanged (Fig. 1d) [31]. Recent work suggested that FL-VD is one of the most common types of VD and might cause more lung damage in patients with ARDS conditions than other types of VD due to the delivery of large tidal volumes [32, 33]. Therefore, in this study, we aim to identify FL-VD breaths against normal breaths, quantify their severity and relate the severity metric with healthcare variables and interventions. For that, we construct new pressure and volume models that can recapitulate the necessary features of FL-VD breaths for a cohort of intensive care unit (ICU) patients. The model estimates are used to identify FL-VD breaths against normal (NL) breaths. We demonstrate that the model provides a powerful continuous severity metric to quantify FL-VD breaths. We also explore the correlation between model-estimated and ventilator-reported parameters to understand the association of FL-VD breaths with healthcare processes.

## METHODS

### Defining clinically relevant features in the pressure and volume ventilator waveform signals

During MV, pressure, volume and flow waveform signals can be used to understand respiratory mechanics, including lung condition and patient-ventilator interaction. To develop a quantitative interpretation of these waveform signals, and to define the hypothesis-driven approach, we break down the waveform into several temporal features and look at how these features vary over breaths and patients. Specific deformations in these features might reflect lung dysfunction and dyssynchronous patient-ventilator interaction in the form of VD when the signal is not controlled via the ventilator [12, 13]. For that, we reviewed extensive human ventilator waveform data, both without and with VD, and identified three temporally ordered processes in the pressure signal (Fig. 2a) and two processes in the volume signal (Fig. 2b). In the pressure waveform, feature P1 is the gradient of the inspiration signal, and it is hypothesized to correspond to the volume-dependent lung compliance when the flow rate is constant [34]. Feature P2 controls the peak and the plateau pressures and how the waveform is deformed during constant pressure, which might reflect patient-ventilator interaction in the FL-VD breaths [35]. Feature P3 corresponds to the gradient of the expiration signal. Additionally, the height of the waveform represents the peak pressure. Note that some clinically relevant features might span between multiple processes or present at the intersection such as at the end of inspiration and the beginning of expiration. In such cases, the features can be characterized over the relevant interval in the breath.

**Figure 2:**
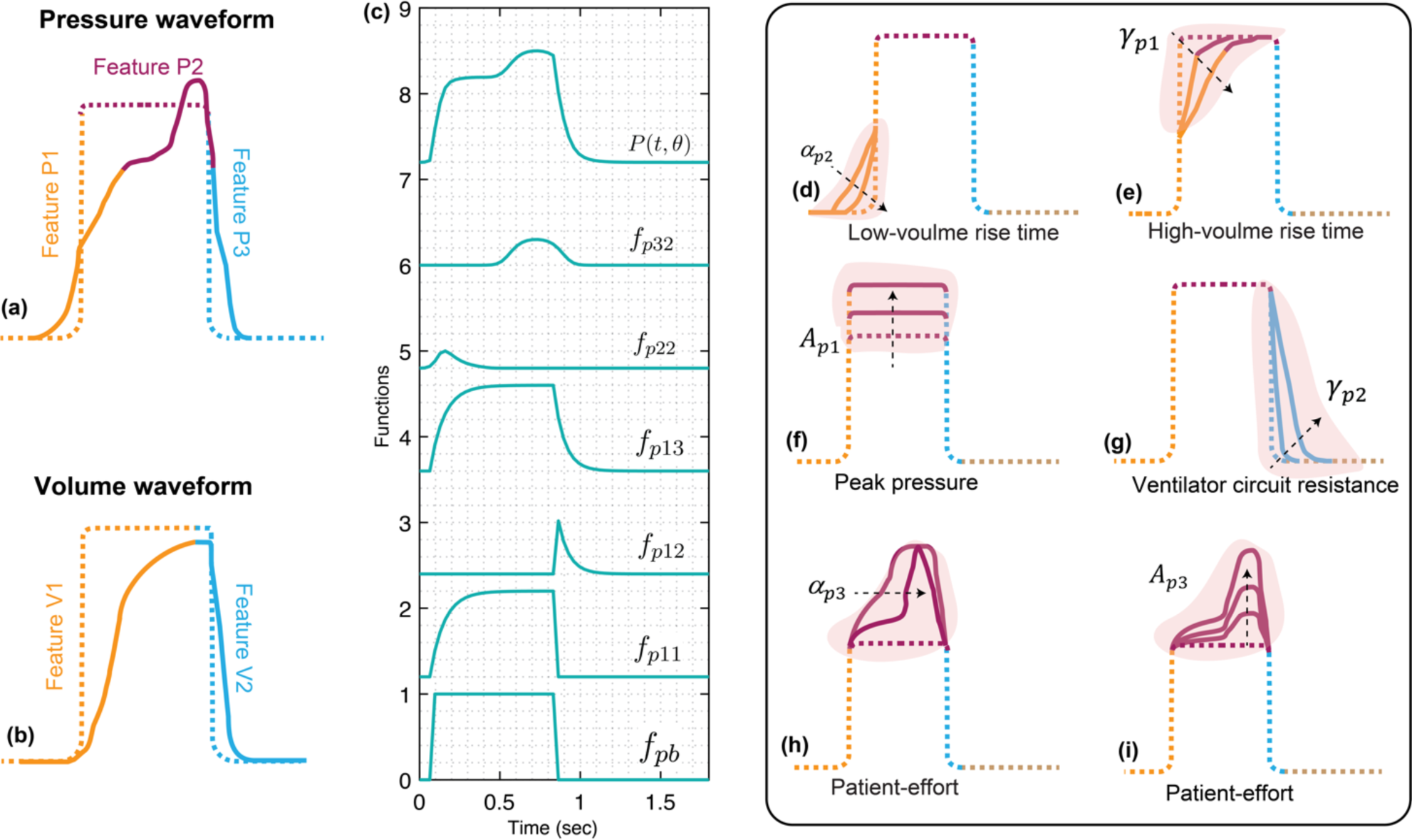
Using the clinically relevant features in the pressure and volume waveforms to construct the VD-deformed lung ventilator model. Graphical representation of theoretical deformed **(a)** pressure and **(b)** volume waveforms. A breath is divided into several temporal processes which are linked with interpretable pathophysiology for qualitatively assessing the lungs state or detecting a VD. The pressure signal has three temporal processes while the volume signal has two temporal processes that might reflect lung condition and patient-ventilator interaction when they are free variables (see Methods). **(c)** Simulated response of various terms that make up the deformed pressure model (P). Equations 1-10 were used to simulate the response within each term with parameter values *θ* = 0.2, *α*_*p*1_= 2 =10^4^, ***β*_*p*1_** = 0.88, **φ_*p*1_** = −0.983, ***γ*_*p*1_**= 3.25, ***γ*_*p*2_**= 2.62, ***A*_*p*1_** = 1, ***CP*** = 0*, α***_*p*2_**= 92.4, ***β*_*p*2_** = 0.984, **φ_*p*2_** = −1.22, ***A*_*p*2_** = 0.2***, α*_*p*3_**= 50, ***β*_*p*3_** = 0.983, **φ_*p*3_** = −0.66, ***A*_*p*3_** = 0.3. Y-axis was normalized to represent all the model components in a sequential manner. **(d-i)** In the pressure signal, the initial gradient of the pressure signal during inspiration at low volume is controlled by **(d)** the *α*_*p*2_ parameter. Higher values result in a slower rise. **(e)** The gradient of the rising signal after the inflection point is controlled by the ***γ*_*p*1_** parameter. Higher values result in a slower rise. **(f)** The amplitude of the waveform can be altered using the parameter ***A*_*p*1_**. **(g)** The gradient of the falling signal during expiration can be modified by the ***γ*_*p*2_** parameter. Higher values result in a slower fall. **(h)** The shape of the peak at the plateau is regulated by the ***α*_*p*3_** parameter such that higher values result in a sharper peak. **(i)** The amplitude of this peak is controlled via the ***A*_*p*1_** parameter. Note that the breaths were drawn to highlight the features which might contain useful information about lung condition and patient-ventilator interaction when the respective variable is free. If the variable is controlled, e.g., volume during VCV, the waveform features represent ventilator settings. Additional interpretations including for the volume model are mentioned in the supplementary material.

In the volume waveform, there are two features A and B as shown in Fig. 2b. Features V1 and V2 are the inspiration and expiration signals hypothesized to correspond to lung compliance and expiratory time constant, respectively. Additionally, the height of the waveform represents the tidal volume. In hybrid ventilation modes, there may be scenarios where both pressure and volume variables are partially controlled. Therefore, in those cases, both waveforms can be confounded in additionally complex ways and require more nuanced interpretation. In this work, we ignored the flow signal as it can be derived from the volume model signal.

### Formulating a deformed lung ventilator model that can capture FL-VD breaths

The basic framework of the model starts with a periodic rectangular waveform and then functional terms are added that produce specific features in the pressure and volume waveform signals individually [30]. In this way, the models track only the features’ specific deviation and not every wiggle encountered in the waveforms. Deformation in these features over several breaths might correspond to time-sensitive lung conditions, while damage-specific deviations can help to understand damage pathophysiology and patient-ventilator interactions. Additionally, in clinical settings, the pressure and volume signals can be strongly or weakly coupled during MV, which is controlled via the mode of ventilation [12]. We, therefore, use respiratory rate only to couple these signals. This approach allows independent modeling of these signals and their respective components such that clinical, physiologic, and ventilator-based knowledge can constrain the model. We named this model the VD-deformed lung ventilator (VD-DLV) model, which contains information about lung physiology and ventilator dynamics. The VD deformed pressure model that may capture the diversity of FL-VD breaths is given by:

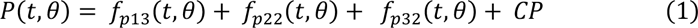

where *θ* is the parameter vector, and *t* is time. The *f_p_*_13_ component is the baseline model that produces an idealized pressure signal. The *f_p_*_22_ component controls the slope near the inflection point or the derivative of the VD-induced deformity, while the *f_p_*_32_ component adds a deformation during or at the end of the inspiration signal that might reflect dyssynchronous patient-ventilator interaction in the form of FL-VD associated features [5]. *CP* is the constant baseline pressure, known as the positive end-expiratory pressure (PEEP) [36]. The baseline model uses a periodic rectangular waveform signal, which is in the form of:

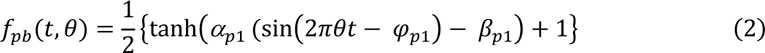

And the baseline model is given by

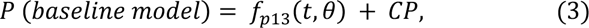

where

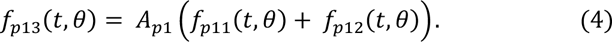

The *f_p_*_13_ component produces the inspiration (Feature P1) and the expiration (Feature P3) part of the pressure signal via the *f_p_*_11_ and *fp*_12_ components, respectively.

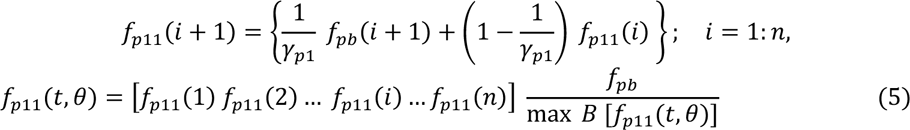

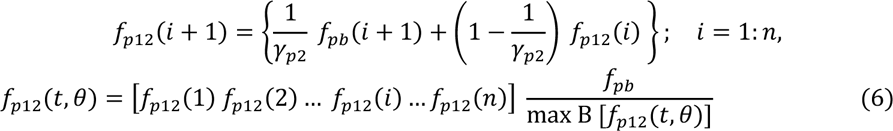

such that *f_p_*_11_(*i* = 1) = *f_p_*_12_(*i* = 1) = 0 and the *i*′*s* are aligned with the corresponding *t*. Here, *B* represents a single breath. The deformities in the waveform at the beginning and during the inspiration signal are produced by the *f*_*p*22_ and the *f_p_*_32_ components respectively, which are defined as:

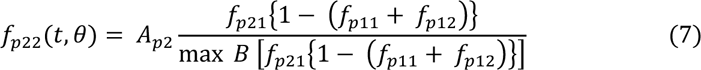

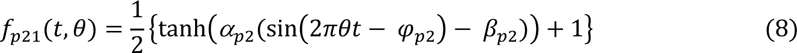

and

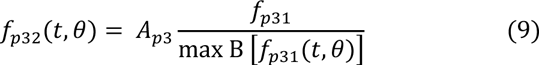

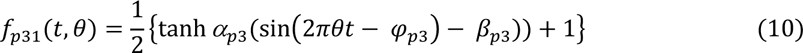

A short description of how the model parameters contribute to the model is provided in Table 1, and how each of the model components contributes to the deformed model is shown in Fig. 2c. Figure 2d-i and Supplementary (SI) Fig. S1 show how some of the important model parameters such as *α_p_*_2_, *γ_p_*_1_*, α*_*p*3_, *A*_*p*1_, and *A_p_*_3_ impact the pressure waveform in a targeted way and have interpretable pathophysiology with hypothesized lung conditions that clinicians often use. The volume model has a similar construction as the pressure model and it is described in section I of the supplementary material along with the relevant plots (SI Figs. S2, S3).

**Table 1:** Interpreting VD deformed lung ventilator pressure model parameters. The parameters that are correlated with known measures of lung physiology are in bold.

| Parameter | Model relevance | Physiological/Clinical relevance (with increased values) |
| --- | --- | --- |
| **Pressure model** |  |  |
| $\theta$ | Number of breaths/sec. Higher values result in a higher number of breaths/sec. | - |
| $\beta_{p1}$ | I:E ratio. Higher values result in a smaller duration of inspiration. | - |
| $\alpha_{p1}$ | Smoothness of the square waveform. Higher values result in a sharper transition. | - |
| $\varphi_{p1}$ | Starting of the $f_{bp}$ function. Higher value results in a more delayed response. | - |
| $\alpha_{p2}, \beta_{p2}$ | Gradient of the pressure signal at low volume. Higher values result in a slower rise. | Higher low volume compliance. |
| $\varphi_{p2}$ | Starting of the inspiration point. Higher value results in a more delayed response. | - |
| $\gamma_{p1}$ | Gradient of the rising signal after inflection point. Higher values result in a slower rising signal. | Higher high-volume compliance. |
| $\gamma_{p2}$ | Gradient of the falling signal. Higher values result in a slower falling signal. | - |
| $\alpha_{p3}, \beta_{p3}, \varphi_{p3}$ | Gradient, width, and position of the bump produced by the $f_{p32}$ component. | |
| $A_{p1}$ | Peak amplitude | Lower overall compliance. |
| $A_{p2}$ | Higher values increase the amplitude of the $f_{p22}$ component. | Moves upper inflection point up |
| $A_{p3}$ | Amplitude of the bump produced by the $f_{p32}$ component | |
| $CP$ | Constant pressure base line value | PEEP |

### Relating the VD-DLV model parameters with the lung function parameters and ventilator dyssynchrony

In our modeling framework, clinical and physiologic knowledge was added to the models’ parameters by allowing observable pressure and volume diversity. Each physiological and VD-associated feature is controlled via individual interpretable parameters. Therefore, we can infer lung condition by qualitatively assessing the effects of parameter changes on these features. Note that these interpretations are valid only when the ventilator settings do not change breath-to-breath and the respective parameters are not controlled by the ventilator.

In the pressure signal, the *α_p_*_2_, *β*_*p*2_, *γ*_*p*1_, and *A_p_*_1_ parameters may reflect aspects of lung compliance when the pressure signal is not controlled via a ventilator. The *α_p_*_2_, *β_p_*_2_ parameters describe low-volume compliance (Fig 2d, SI Fig. S1), while the parameter *γ_p_*_1_ correlates with compliance at higher volumes in the breath (Fig 2d, SI Fig. S1). The *A_p_*_1_ parameter defines the peak pressure and is inversely correlated with compliance (Fig 2e, SI Fig. S1). The *γ_p_*_2_ parameter might be used to account for ventilator circuit resistance (Fig 2g, SI Fig. S1). Additionally, some model parameters can be used to produce VD-associated specific features. As the scope of this paper is limited to NL and FL-VD breaths, we focus on the deformity produced by the *f*_*p*32_ component during or at the end of inspiration. This deformity, which is often reflected by scoping in the pressure signal (Fig. 1d), is controlled by the *α_p_*_3_, *β_p_*_3_, *A_p_*_3_ and *Ap*_3_ parameters (Fig 2h,I, and SI Fig. S1) [15]. A similar interpretation of the volume model is provided in section I of the supplementary material.

### Data collection and annotation

We included patients admitted to the University of Colorado Medical, Surgical, Neurosurgical, and COVID ICUs. Patients between 18 and 89 years old, needing mechanical ventilation, and with the ARDS by the Berlin definition or ARDS risk factors were enrolled within 24 hours of intubation [2]. Alternatively, patients could be enrolled within 24 hours of the placement of an esophageal balloon by the clinical team. At-risk patients were defined as intubated patients with a mechanism of lung injury known to cause ARDS, who have not yet met chest x-ray or oxygenation criteria for ARDS. All patients were ventilated with a Hamilton G5 ventilator. IRB approval was obtained for this study. Patients were excluded if: a) less than 18 years of age, b) pregnant, c) imprisoned, d) with esophageal injury, recent esophageal or gastric surgery (3 months), e) tracheal-esophageal fistula, f) facial fracture, or g) active or recent (3 months) variceal bleed or banding.

Patient characteristics were extracted from the electronic health records (EHR). Continuous ventilator data were collected using a laptop connected to the ventilator using Hamilton DataLogger software (Hamilton, v5.0, 2011) to obtain time-stamped measures of airway pressure (paw), flow, volume, and esophageal pressure measurements (pes). Data were recorded at 32ms intervals. Additionally, the DataLogger software collects ventilator mode and ventilator settings. Data were collected at the time of enrollment for up to 48 hours after esophageal balloon placement to reduce the effects of length of time bias. An esophageal balloon pressure monitor (CooperSurgical; Truball, CT) was inserted into the sedated and intubated patient at the time of enrollment for all patients. The placement was confirmed by the balloon occlusion test [37]. The Colorado Multiple Institutional Review Board approved this study and waived the need for informed consent.

Two types of breaths were identified for this study (Table 2) while other types were grouped into families of VD to simplify identification. First, the family of normal appearing breaths is characterized by an appearance of the airway pressure, flow, and esophageal pressure waveforms of a patient breathing on the ventilator without respiratory muscle contraction deforming the expected airway pressure and flow waveforms. Normal-appearing breaths can be passive (ventilator driven) or spontaneous (initiated by the patient). Second, the family of flow-limited (FL) breaths is defined by patient effort (a negative pes deflection during inspiration) exceeding the inspiratory flow achieved by the ventilator. The results in a coving of the airway pressure waveform (Fig. 1c). Finally, the remainder of the breaths were categorized as miscellaneous including all other types of VD breaths.

**Table 2:**
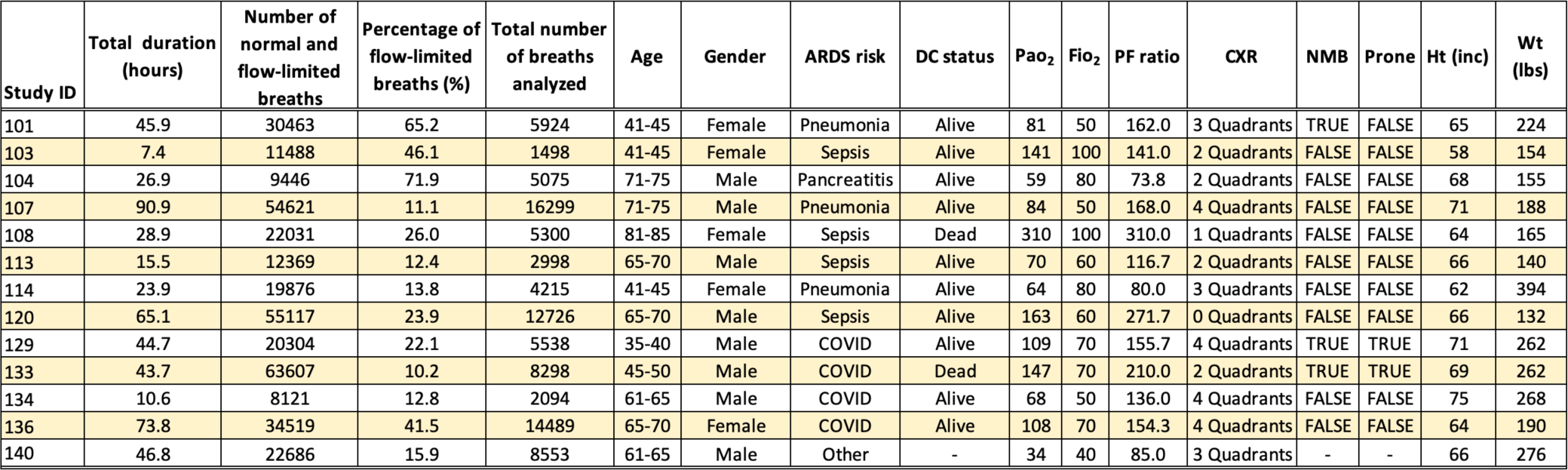
Baseline patient information. Each patient was assigned a random study ID. An in-house developed ML algorithm was used to annotate normal and flow-limited breaths (column 3) from the continuous ventilator data while ignoring other VD breaths such as double-triggered, auto-triggered, etc. [5, 15]. In this study, we considered and analyzed patients with more than 10% flow-limited breaths (column 4). The total number of breaths analyzed for each patient is shown in column 5 (see Methods). Patient 102 was ignored as only 70 breaths in total were available to annotate.

| Study ID | Total duration (hours) | Number of normal and flow-limited breaths | Percentage of flow-limited breaths (%) | Total number of breaths analyzed | Age | Gender | ARDS risk | DC status | Pao<sub>2</sub> | Fio<sub>2</sub> | PF ratio | CXR | NMB | Prone | Ht (inc) | Wt (lbs) |
| --- | --- | --- | --- | --- | --- | --- | --- | --- | --- | --- | --- | --- | --- | --- | --- | --- |
| 101 | 45.9 | 30463 | 65.2 | 5924 | 41-45 | Female | Pneumonia | Alive | 81 | 50 | 162.0 | 3 Quadrants | TRUE | FALSE | 65 | 224 |
| 103 | 7.4 | 11488 | 46.1 | 1498 | 41-45 | Female | Sepsis | Alive | 141 | 100 | 141.0 | 2 Quadrants | FALSE | FALSE | 58 | 154 |
| 104 | 26.9 | 9446 | 71.9 | 5075 | 71-75 | Male | Pancreatitis | Alive | 59 | 80 | 73.8 | 2 Quadrants | FALSE | FALSE | 68 | 155 |
| 107 | 90.9 | 54621 | 11.1 | 16299 | 71-75 | Male | Pneumonia | Alive | 84 | 50 | 168.0 | 4 Quadrants | FALSE | FALSE | 71 | 188 |
| 108 | 28.9 | 22031 | 26.0 | 5300 | 81-85 | Female | Sepsis | Dead | 310 | 100 | 310.0 | 1 Quadrants | FALSE | FALSE | 64 | 165 |
| 113 | 15.5 | 12369 | 12.4 | 2998 | 65-70 | Male | Sepsis | Alive | 70 | 60 | 116.7 | 2 Quadrants | FALSE | FALSE | 66 | 140 |
| 114 | 23.9 | 19876 | 13.8 | 4215 | 41-45 | Female | Pneumonia | Alive | 64 | 80 | 80.0 | 3 Quadrants | FALSE | FALSE | 62 | 394 |
| 120 | 65.1 | 55117 | 23.9 | 12726 | 65-70 | Male | Sepsis | Alive | 163 | 60 | 271.7 | 0 Quadrants | FALSE | FALSE | 66 | 132 |
| 129 | 44.7 | 20304 | 22.1 | 5538 | 35-40 | Male | COVID | Alive | 109 | 70 | 155.7 | 4 Quadrants | TRUE | TRUE | 71 | 262 |
| 133 | 43.7 | 63607 | 10.2 | 8298 | 45-50 | Male | COVID | Dead | 147 | 70 | 210.0 | 2 Quadrants | TRUE | TRUE | 69 | 262 |
| 134 | 10.6 | 8121 | 12.8 | 2094 | 61-65 | Male | COVID | Alive | 68 | 50 | 136.0 | 4 Quadrants | FALSE | FALSE | 75 | 268 |
| 136 | 73.8 | 34519 | 41.5 | 14489 | 65-70 | Female | COVID | Alive | 108 | 70 | 154.3 | 4 Quadrants | FALSE | FALSE | 64 | 190 |
| 140 | 46.8 | 22686 | 15.9 | 8553 | 61-65 | Male | Other | - | 34 | 40 | 85.0 | 3 Quadrants | - | - | 66 | 276 |

A training set of 10,000 breaths from 30 patients was randomly selected for manual review and labeling by one of the investigators (PDS). The manual review was used as the gold standard to identify FL-VD [4]. Algorithm development followed the classic supervised-learning classification problem. The process of developing this classification model follows five steps: 1) data cleaning and verification, 2) feature engineering, 3) feature selection, 4) model for training, and 5) model validation.

### Data selection

Raw ventilator waveform data typically includes 25,000 to 35,000 breaths per day per ICU patient, which can be too large to estimate. Accordingly, we devised a systematic framework to create a reduced dataset for a cohort of patients that is more manageable to estimate and can capture the variations in the lung state during prolonged MV. In the current study, we only consider NL and FL breaths while ignoring other types of VD breaths. The amount of FL breaths available in a dataset can vary significantly from one patient to another and even in the same patient over time. This variability is normal in clinical data as the VD types and frequency are highly dependent on many healthcare variables and interventions. We, therefore, created filtered datasets for each patient that allow us to conduct this steady in a meaningful way. To create this filtered dataset, we used two criteria.

First, because our analysis focuses on FL-VD, patients with 10% or more FL breaths in the originally recorded dataset were considered. Each breath in the originally recorded dataset was annotated using the ML method described above in the data selection and annotation section. We then determined the percentage of each type of breath out of all the recorded breaths. Out of 62 patients, 30 patients’ data were available to annotate at the time of the study, and of those 13 met our first criterion. For each of these 13 patients, filtered datasets were created with only NL and FL breaths (Table 2). Each patient was assigned a random identifier (study ID) for the purpose of this paper. This study ID was not known to anyone outside the research group.

Second, to manage the computational cost, we randomly selected 100 breaths in a 30 min sampled window using the RAND function in MATLAB. This procedure was repeated for 30 min windows covering the entire duration of the ventilator waveform recording. We expected to have ∼600 breaths per 30 min window if only NL and FL breaths were present in filtered datasets. However, the total number of NL and FL breaths varies in the sampled time window depending on the types of breaths present in the original datasets, which are subjected to the patient’s condition, ventilator settings and healthcare interventions. For example, some patients might have FL-VD breaths distributed throughout the time during MV while others might have them for a particular time duration only. Due to this variability in the number of breaths over time, we observed three different cases. In case 1, where the total number of NL and FL breaths was more than 100 in the sampled time window, a maximum of 100 breaths were selected (SI Fig. S4). In case 2, less than 100 combined NL and FL breaths were present in the sampled time window, and the maximum number of available breaths was taken, ranging from 1 to 100 (SI Fig. S5). In case 3, no NL or FL breaths were present in a 30 min time widow, and no breaths were selected (SI Fig. S6). Depending on the patient, some cases dominated others. The total number of breaths analyzed for each patient is shown in column 5 of Table 2.

### Data estimation and analysis

We partition the ventilator waveform data by breath, labeled with index *q*. Each breath, *B_q_*, which is measured with 32.5 samples per second, had N points per breath with index *i*. By doing this, feature-specific variation over breaths can be tracked easily by relating the estimated parameters per breath. The model estimation is computed as a smoothing problem using a gradient-based minimization algorithm for nonlinear functions using the MATLAB FMINCON function [38-40]. To ensure a robust solution and quantify uncertainty, we also used the MATLAB MULTISTART function, which performs the optimization using multiple starting points selected from realistic lower and upper bound values (constraints) using an iterative method. These bounds also define the constraints employed by the parameter estimation problem in the optimization scheme [41, 42]. The VD-DLV model includes two state variables, pressure and volume with one overlapping parameter, the respiratory rate (*θ*), which was fixed. The estimation of the pressure model parameter is more complex as it has fifteen parameters (*α*_*p*1_, *β*_*p*1_, *A*_*p*1_, *γ*_*p*1_, *γ*_*p*2_, *A*_*p*1_, *CP*, *α*_*p*2_, *β*_*p*2_, *A*_*p*2_, *A*_*p*2_, α_*p*3_, *β*_*p*3_, *A*_*p*3_, *A*_*p*3_). These parameters contribute to the specific features in the pressure waveform, allowing the parameter to be estimated relative to the specific, time-limited feature.

Estimating model parameters is relatively straightforward when the model is identifiable means there is a unique solution in terms of best parameter values for a given data. Even though the VD-DLV model is constructed such that most of the model parameters are active temporally and not during the entire single breath, non-uniqueness of non-convergence of solutions can still occur [43-45]. To minimize such cases, two approaches were used. In the first approach, all model parameter ranges were generally constrained to lie within physiologically possible values.

In the second approach, we gradually add deformations to the model allowing us to estimate the parameters that correctly represent the associated features. For that, first the baseline model (Eqn 3) was used to estimate the parameters that produce the prominent features in the pressure waveform signal such as I:E ratio, amplitude, inspiration and expiration slopes and PEEP without any FL-VD associated deformities. We then use these estimates as initial parameter values while using the VD-DLV model (Eqn 1), which produces all the features in the waveform data (see Methods). These approaches were implemented using an iterative method where the estimation of the first run was used as an input for the next run with tighter constraints. In particular, for a given breath *B_q_*, the pressure signal of every breath was estimated at least two times and up to five times, denoted as *Run 1* to *5* as shown in Fig. 3a.

**Figure 3:**
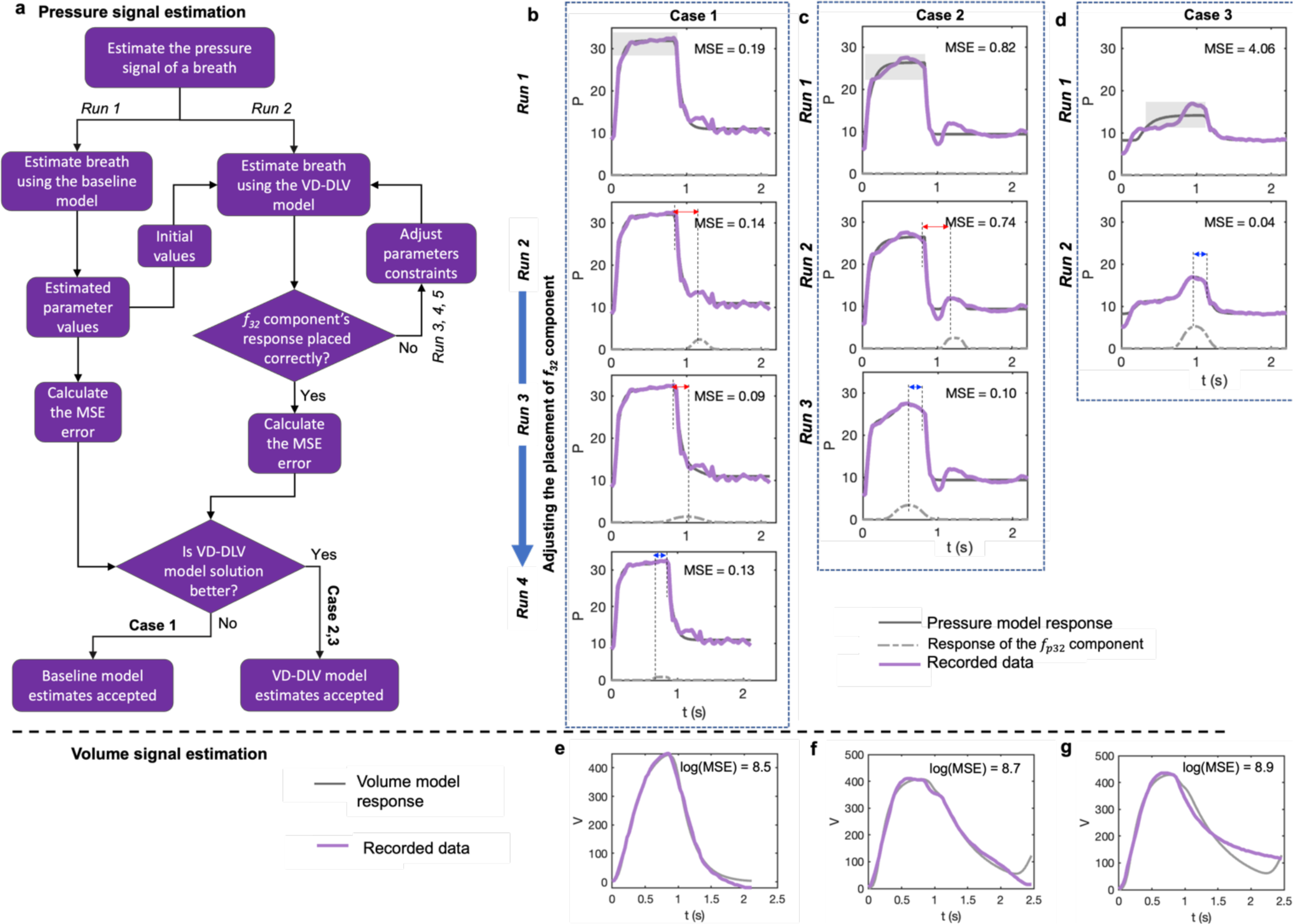
The VD-deformed lung ventilator (VD-DLV) model can accurately reproduce all the essential features found in pressure and volume waveform signals of the FL-VD breaths. **(a)** Flow chart of the estimation methodology for the pressure waveform signal. The pressure signal baseline model comprises the *f*_*p*1#_ component and *CP*, while the VD-DLV model comprises all three components (*f*_*p*1#_, *f*_*p*2$_, and *f*_*p*32_) and *CP*. While using the VD-DLV model for estimation, the *A*_*p*3_ parameter constraints were adjusted to ensure that the deformation produced by the *f*_*p*32_ component is placed during before the end of inspiration (see Methods). The final parameters estimations were accepted either from the baseline or the VD-DLV model based on the Mean Squares Errors (MSE) score produced by these models (see Methods). **(b-d)** The pressure waveforms of three representative breaths where the final model’s response is shown in sold-grey lines, while the recorded data is shown in solid-colored lines. In Cases 1 and 2, after *Run 2*, the *A*_*p*3_ parameter constraints were adjusted as the response of the *f*_*p*32_ component was placed incorrectly-shown by the red arrow in *Run 2*. The same process was repeated for Case 1 in *Run 3* and *4* while for Case 2 a positive difference was achieved in *Run 3*-shown by the blue arrow. By this approach, we found the optimum solution for Cases 1, 2 and 3 in *Run 4*, *3* and *2*, respectively. However, for Case 1, unlike Cases 2 and 3, the VD-DLV model did not significantly reduce the MSE, and hence the solution of the baseline model was accepted. For MSE calculation, the regions highlighted in the plots of *Run 1* for each case were considered (see Method). **(e-f)** The estimated volume waveform signal for **(e)** Case 1, **(f)** Case 2, and **(g)** Case 3. The estimated model parameters for each Case and *Run* are shown in SI Table S3-S5.

In *Run 1*, we estimated only the baseline model (Eqn 3) parameters (*α*_*p*1_, *β*_*p*1_, *A*_*p*1_, *γ*_*p*1_, *γ*_*p*2_, *A*_*p*1_, *CP*). From *Run 2* onwards, VD-DLV model parameters (*α*_*p*1_, *A*_*p*1_, *γ*_*p*1_, *A*_*p*1_, *α*_*p*2_, *β*_*p*2_, *A*_*p*2_, *A*_*p*2_, α_*p*3_, *β*_*p*3_, *A*_*p*3_, and *A*_*p*3_) were estimated. The values of *β*_*p*1_, *γ*_*p*2_, and *CP*, were fixed from *Run 2* onwards to maintain the characteristic shape of the waveform such as the I:E ratio and PEEP, which are unaffected by VD. The estimates of *Run 1* were used as initial parameter values for *Run 2*; similarly, in subsequent runs, estimations of the previous *Run* were used as initial parameter values. After *Run 2*, estimation was stopped if the deformity produced by the *f*_*p*32_ component was placed before the end of the inspiration signal – a characteristic of FL-VD breaths [32]. This was tracked by determining the difference between the time the inspiration signal ended and the time the *f*_*p*32_ component achieved maximum amplitude. A negative difference means the component’s response was placed after the inspiration part of the breath, which is incorrect for FL-VD; hence, additional *Runs* were necessary. Subsequently, from *Run 3 onwards*, the parameters were estimated again with adjusted constraint values for the *A*_*p*3_ parameter while keeping the constraints of other parameters the same. During the adjustment, the upper bound of the *A*_*p*3_ parameter was modified by the algorithm (Table S2) that shifted the *f*_*p*32_ component’s response towards the inspiration part of the breath. The same process was repeated in *Runs* 4 and 5 if the *f*_*p*32_ component’s response was placed after the end of inspiration.

The final parameter estimation was accepted either from the baseline model (*Run 1*) or from the VD-DLV model (*Run 2-5*) based on the Mean Squared Errors (MSE) scores. For that, given the FL-VD associated deformities arises only in the feature P2 part of the pressure signal, all the data points that satisfied *f*_*p*11_ ≥ 0.75 ξ *A*_*p*1_ before the end of the inspiration were considered. Those regions are highlighted in grey in the plots of *Run 1* for each Case in Fig 3b-d. For a given breath *B_q_*, estimates of the VD-DLV model were accepted only if 1) the MSE produced by the VD-DLV model is 50% or lesser than the MSE produced by the baseline model, and 2) MSE produced by the baseline model from *Run 1* is more than the mean of MSE for all the estimated breaths by the VD-DLV model for a given patient. Otherwise, the baseline model estimates from *Run 1* were accepted as an optimum solution. Each *Run* comprises 100 iterations with different initial guesses to quantify parameter uncertainty and find global or multiple minima depending on the solution surface for each parameter [42]. Details for volume model estimation are provided in the Supplementary material.

## RESULTS

### VD-DLV model accurately captures diverse features produced by FL-VD breaths

Our primary objective is the quantification of FL-VD breath incidence and severity. This requires estimating FL-VD breaths associated features in the pressure waveform and then using those model estimates to define a severity metric to phenotype FL-VD breaths. In the deformed model, the *f*_*p*32_ component is designed to track the characteristic features of FL-VD breaths. However, there may be other deformations in the breath where this component response might lead to optimum fit, i.e., producing the lowest error but having no physiological meaning. To minimize such cases, we have developed an iterative approach to correctly estimate the FL-VD associated features (Fig. 3a). In this approach, the baseline model estimates were used to constrain the VD-DLV model solution and based on the improvement on the MSE score, the solution of the VD-DLV model or the baseline model based was accepted (see Methods).

We tested this approach on patient ID 104 where we sampled 5,075 breaths from the filtered dataset (see Method) and then performed parameters estimation breath-by-breath. Figure 3b-d shows three representative breaths (solid-colored lines) along with the model response (sold-grey lines) at the optimum parameter values (SI Table S3). We found that in the first two representative breaths (Cases 1 and 2), the response of the *f*_*p*32_ component was not correctly placed as it was meant to track the deformity during or at the end of inspiration (shown in the blue arrow) and should not be during the expiration (shown in the red arrow). Therefore, additional *Runs* were necessary for Cases 1 and 2 where the *Φ*_*p*3_ parameter constraints were adjusted automatically by the algorithm. The optimum fits were found in *Runs 4* and *3*, respectively. Finally, unlike Case 1, in Cases 2 and 3, the final estimates significantly reduced MSE, and so the deformed model’s solutions were accepted as optimum solutions (see Methods). The parameter estimates and the respective model response for each Case and *Run* are shown in SI Tables S4-S6 and SI Figs. S7-S9, respectively. The estimation of the volume signal of each representative breath is shown in Fig. 3e-g.

### Computing FL-VD breaths phenotype using the VD-DLV model estimate-defined VD severity metric

The VD-DLV model is a feature-specific model, where variations in the model parameters produce deviations in specific waveform features. As such, we can develop a systemic framework to phenotype the severity of FL-VD breaths by correlating variation in the feature-specific parameters. For patient ID 104, all the estimated breaths (5,075) are shown in the order they appeared in the recorded dataset, starting with the first estimated breath, which is assigned t=0 (Fig. 4a). The plot of the mean squared error (MSE) shows that the VD-DLV model provides an accurate estimate of all breaths (Fig. 4b).

**Figure 4:**
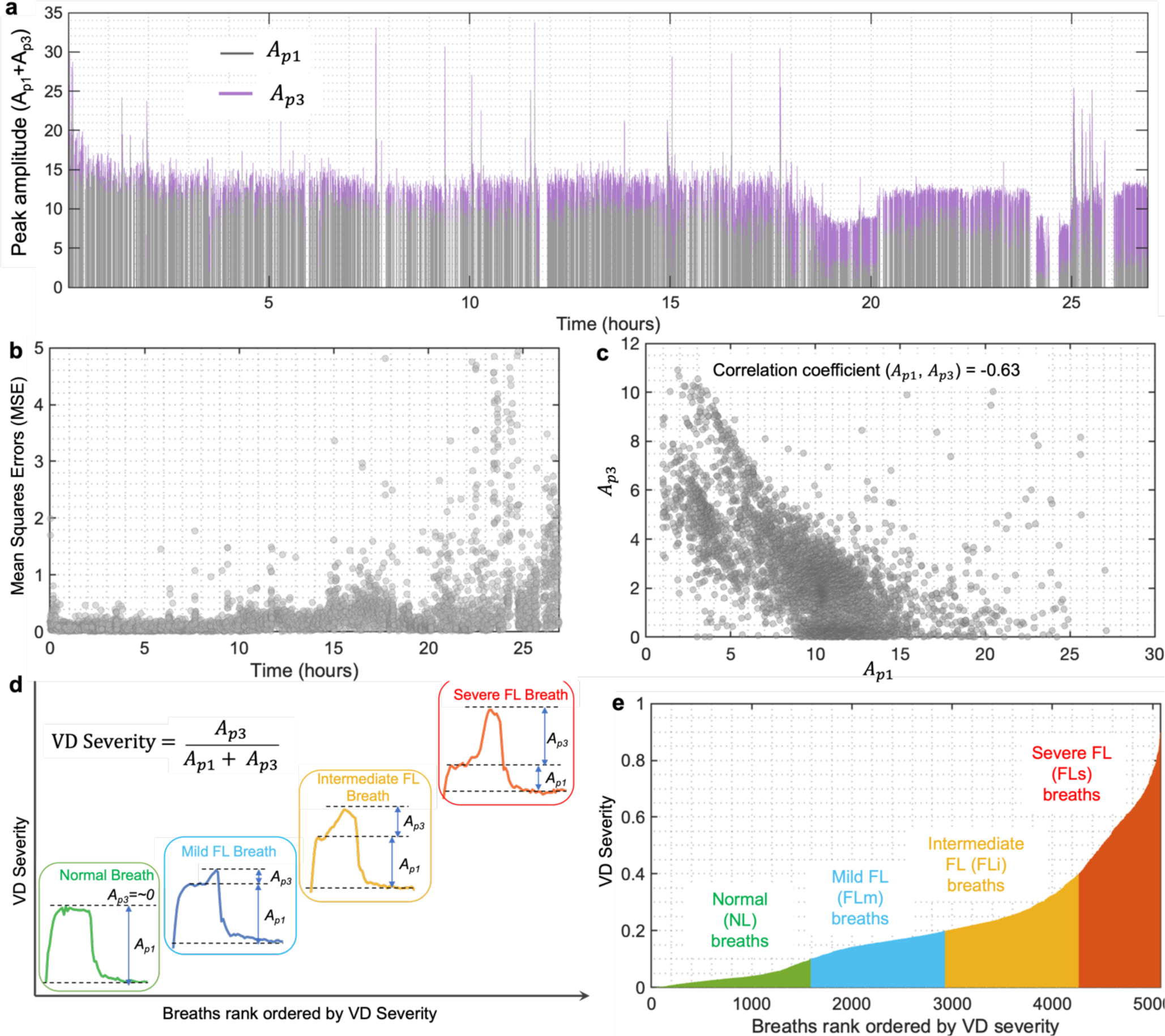
Identifying and quantifying FL-VD breaths using the VD-DLV model-defined VD severity metric. **(a)** For patient ID 104, all the estimated breaths (5,075) are shown as they appeared in the recorded dataset, starting with the first breath at t=0 hour. A single breath is represented by a bar where the height of the bar corresponds to the respective values of *A*_*p*1_ and *A*_*p*3_ parameters, shown in grey and violet colors, respectively. The *A*_*p*1_ and *A*_*p*3_ parameters control the height of the base waveform produced by the *f*_*p*1#_ component and the height of the bump produced by the *f*_*p*32_ component, respectively. **(b)** Mean squares errors (MSE) for all the estimated breaths. The mean value of MSE and the corresponding standard deviation are 0.3 and 0.53, respectively. (**c)** A negative correlation is observed between the *A*_*p*1_ and *A*_*p*3_ parameters, such that breaths with lower values of *A*_*p*1_ tend to have higher values of the *A*_*p*3_ parameter. **(d)** The VD severity metric tracks the severity of the scalloped-out portion of the pressure waveform, a characteristic feature of FL-VD breaths during a patient-triggered volume breath [32]. **(e)** Quantifying the severity of FL-VD breaths against NL breaths. All the breaths with VD severity<0.1 were classified as NL breaths. All the breaths with 0.25>VD severity>0.1 were classified as mild flow limited (FLm) breaths. All the breaths with 0.4>VD severity>0.25 were classified as intermediate flow limited (FLi) breaths. All the breaths with VD severity>0.4 were classified as severe flow limited (FLs) breaths. Breaths are ordered with respect to the normalized values of *A*_*p*3_, which is *A*_*p*3_/(*A*_*p*1_ + *A*_*p*3_).

One of our hypotheses for FL-VD breath is that the amplitude of the VD deformity determines the severity of dyssynchronous patient-ventilator interaction. To formulate this hypothesis, we focus on the amplitude (*A*_*p*1_) of the baseline model produced versus the amplitude (*A*_*p*3_) of the deformity created by the *f*_*p*32_ component. These two amplitude parameters are strongly inversely correlated, as shown in Fig. 4c. With increasingly severe FL-VD breaths, the airway pressure waveform becomes ‘scooped’ to smaller pressure values. Therefore, we define severity as *A*_*p*3_/(*A*_*p*1_ + *A*_*p*3_), which tracks the FL-VD deformity with respect to the baseline peak pressure value (Fig. 4d). A higher VD severity value means the higher height of the dished-out portion with respect to the height of the base waveform. This means that the more severe breaths will show up at the right side of the plot when the breaths are ranked order by VD severity as shown in Fig. 4d. This normalized value allows generalizing the VD severity metric across many patients as it is independent of the absolute values of the *A*_*p*1_ and *A*_*p*3_ parameters.

The dished-out portion in the pressure signal of a breath might be associated with the inspiratory efforts when the flow is insufficient, causing the pressure signal to scoop out during the inspiratory phase [4]. Therefore, the VD severity metric could be tested clinically to evaluate the patient effort and the severity of FL-VD breaths. We, therefore, use the VD severity metric to determine the presence of FL-VD breaths and rank-order them by severity. For example, breaths with low values of the VD severity metric can be considered NL breaths. Similarly, breaths with relatively high values of the VD severity metric could be considered severe FL-VD breaths. To determine these bounds, we relied on an expert opinion (P.D.S.). However, note that this classification is somewhat hypothetical and might vary person-to-person. We opted to classify the FL-VD breaths into one of three categories-mild, intermediate, and severe while considering NL breaths as a reference. For patient ID 104, this leads to 1591 NL breaths and 1960, 713, and 811 mild (FLm), intermediate (FLi), and severe flow limited (FLs) breaths, respectively, as shown in Fig. 4e.

### VD severity metric can be generalized to phenotype FL-VD breaths for a cohort of ICU patients

We now apply the VD severity metric classification to an independent cohort of patients to test its generalizability. Such generalizability would allow producing accurate results and prevent failures when deploy in practice. For that, using the estimation methodology described in Fig. 3, we estimated individual breaths from the filtered waveform datasets for 12 additional patients who met inclusion criteria (see Methods). The patient characteristics are summarized in Table 2. The corresponding FL-VD severity metric ranked order by severity, of each patient is shown in Fig. 5a. Of 93,007 breaths, over 96% of breaths have MSE<0.5, demonstrating that the estimation methodology can reproduce the diverse features present in the waveform signals for a large and heterogeneous waveform dataset (Fig. 5b). MSE for the volume waveform signals are shown in SI Fig S10.

**Figure 5:**
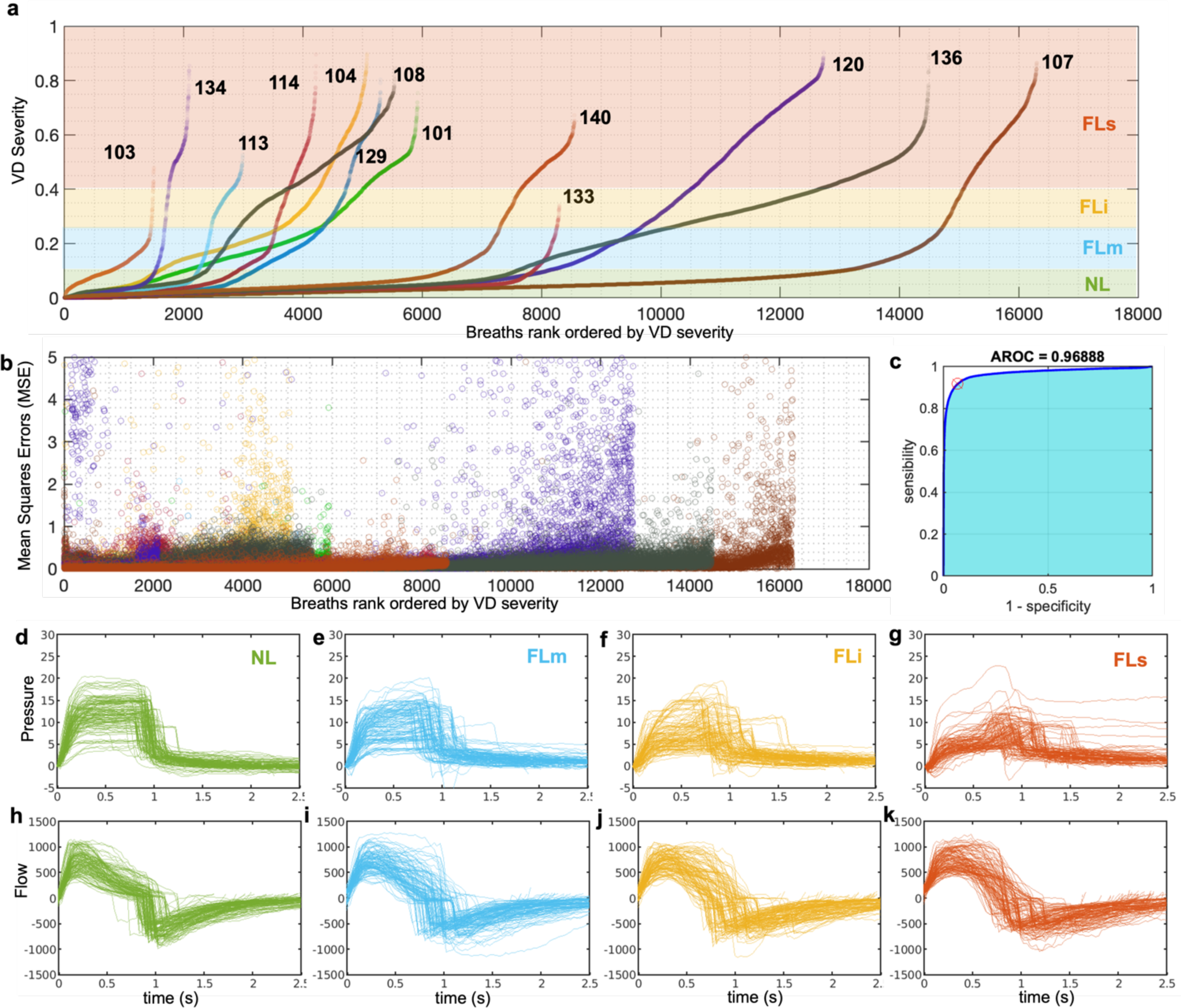
VD severity metric can be generalized to accurately phenotype the severity of FL breaths for a cohort of patients. **(a)** Estimated values of VD severity metric for 13 patients, including patient ID 104. The total number of estimated breaths shown here is 93,007. Each breath is represented by a dot and classified using the VD severity criteria (Fig. 4). **(b)** Mean squares errors (MSE) for all the estimated breaths. The mean value of MSE and the corresponding standard deviation are 0.12 and 0.7, respectively. The total number of 227 breaths was ignored with MSE>5. **(c)** Comparing the VD-DLV model classification of NL and FL-VD breaths with the human-guided ML algorithm (see Methods). An area under the receiver operator curve is 0.97. **(d-j)** Manual validation of phenotypic characterization of FL-VD breaths. Selected **(d-g)** pressure and **(h-k)** flow waveform signals of NL, FLm, FLi, and FLs breaths. Ten breaths were randomly selected from each phenotype for each patient. Since patient ID 133 had no severe FL-VD (FLs) breaths, 130 NL, FLm, and FLi breaths while 120 FLs breaths each are shown here, which were taken from a total of 13 patients. An accuracy of over 98% was achieved.

To cross-validate the accuracy of the VD severity metric, we compared our results with the results obtained from the human-guided ML algorithm (see Methods) and an area under the receiver operator curve (AU-ROC) of 0.97 was achieved [4]. This ML algorithm was already tested in our earlier work with cross-validation and the separate validation set with AU-ROC of over 0.89 for FL-VD breaths compared to the expert manual demarcation [32]. Note that the ML method uses qualitative implementation to differentiate between NL and FL breaths, unlike the VD severity metric, which uses quantitative measurements by accurately taking FL-VD associated features. An additional comparison was carried out to account for the imbalance between the total number of NL and FL breaths using the precision-recall curve and the area under the precision-recall curve of 0.92 was observed (SI Fig. S11).

We also carried out a separate validation to determine how accurately FL-VD-induced features were represented by the VD severity metric. A correct response of the VD-DLV model at the optimum values should reproduce the FL-VD associated features in the pressure waveform signal correctly. For that, we manually went through 510 breaths selected randomly from all the patients. We found that for over 98% of breaths, the phenotypic classification based on the VD severity metric was agreeing with FL-VD associated features present in the pressure waveform signal. The pressure and flow signals of all the selected breaths for each phenotype are shown in Fig. 5d-g and Fig. 5h-k, respectively. The model response for some of the selected breaths are shown in SI Fig. S12-15. Therefore, our FL-VD severity metric can accurately identify the presence and severity of deviations from normal in a way that has a physiologically based hypothesis attached to it.

### Correlating FL-VD severity classification with model and ventilator parameters

The VD severity metric can be used to understand how the severity of FL-VD breaths is associated with waveform features, which can then be used to understand the causes and effects of FL-VD breaths. This severity characterization of breaths (NL, FLm, FLi or FLs) for each patient is shown in Fig. 6a. We used this information to compute the linear correlation coefficient (cc) between the physiologically relevant model parameters and the VD severity metric. An average value correlation coefficient (cc) across all the patients is shown in Table 3. A positive cc value means breaths with higher values of VD severity (such as FLs) will have higher values of the respective parameter, while a negative cc value suggests the opposite. Since any change in the model parameters might be induced by the dyssynchrony itself and might not reflect lung conditions or ventilator settings, a holistic approach must be taken while interpreting these results. For example, the gradient of the rising signal after the inflection point is controlled by the *γ*_*p*1_ parameter. Higher values result in a slower rise and might be referred to as higher high-volume compliance when there is no patient effort. A positive correlation for the *γ*_*p*1_ parameter suggests that FLs breaths take much longer to reach the same pressure value than NL breaths. Similarly, a positive cc value for *the α*_*p*3_ parameters suggests a sharper increase in the pressure signal during the bump at end-inspiration, which is prominent in the FL-VD breaths [5]. Moreover, a positive cc value for the *A*_*v*1_ parameters suggests that the patient takes relatively higher tidal volume in the FLs breaths than NL breaths, as we have previously shown in ML-based analysis [32]. The flow-limited dyssynchrony likely induces all these changes in the pressure and volume signals because of the patient’s effort and not due to changing lung conditions [46]. Therefore, we cannot use the model parameters to assess lung conditions but rather additional markers to identify and quantify FL-VD breaths.

**Figure 6:**
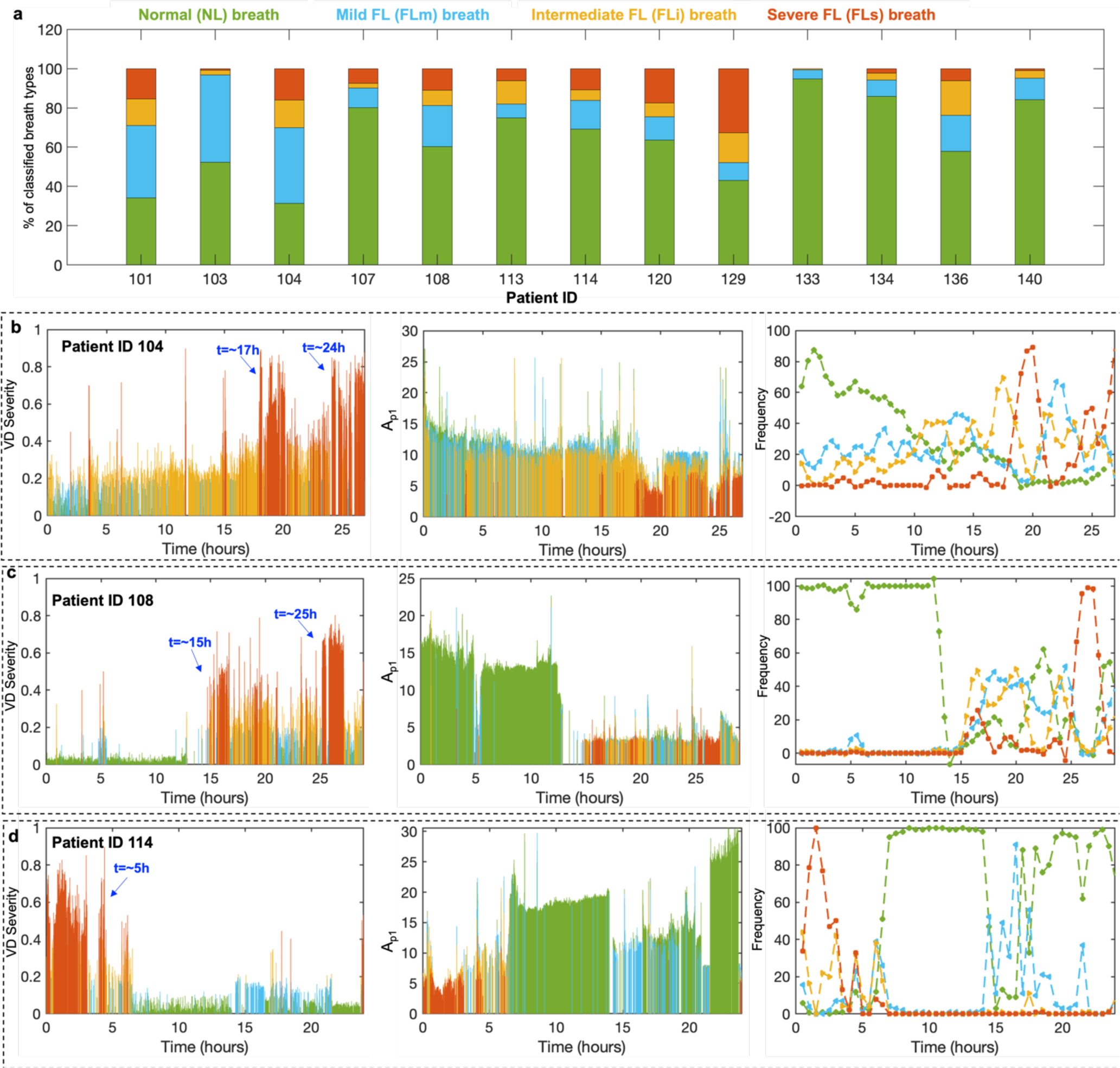
Using the flow-limited VD severity metric to determine observable impact of interventions on the patient state. **(a)** Phenotypic characterization of FL-VD breaths for a cohort of patients using the VD-DLV model defined VD severity metric. **(b-g)** For patient ID **(b)** 104, **(c)** 108, and **(d)** 114, all the estimated breaths are shown as they appeared in the recorded dataset, starting with the first breath at t=0 hour. Each breath is represented using the value of VD severity in bar height, while the color represents the phenotype. The respective values of *A*_*p*1_ parameters of each breath, and the frequency of each phenotype of breaths averaged over 10 min time window are shown. The frequency trajectory of each phenotype was smoothed using the loess function in MATLAB with a 10% overlap.

**Table 3:**
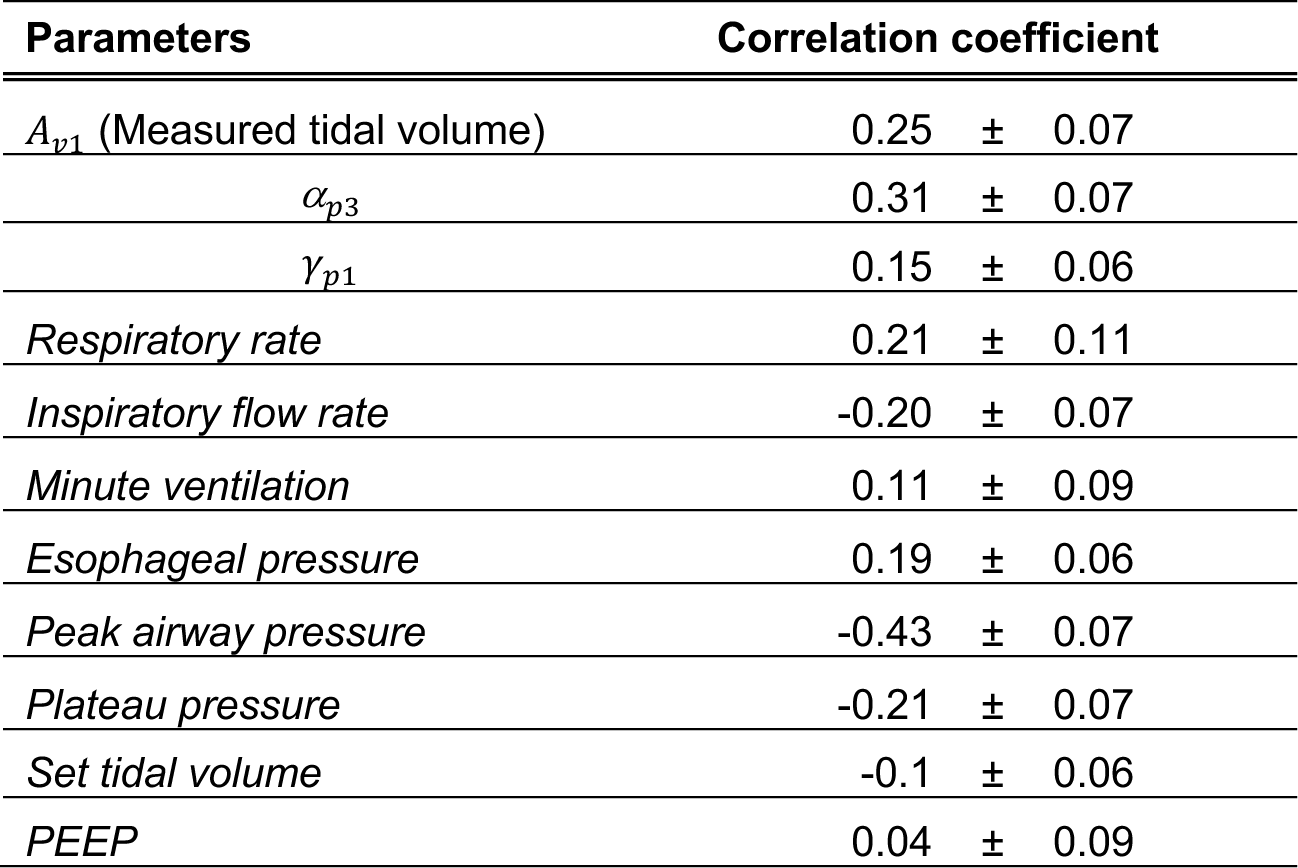
An average value of the linear correlation coefficient across all the patients is shown here. All the parameters highlighted in bold are the VD-DLV model parameters while the MV-measured parameters are italicized. The normalized correlation coefficient values are reported at zero lag between the VD severity metric and either the model parameters or the parameters measured via the ventilator.

We can also determine how ventilator settings relate to the severity of FL-VD breaths, which might help to identify if certain settings lead to more severe FL-VD breaths. For that, we calculated cc values between the ventilator-measured parameter during MV and the VD severity metric (Table 3). As expected, a negative cc value for the flow rate is associated with FL-VD breaths [46]. Likewise, there is a negative correlation between peak airway pressure and FL-VD due to the inspiratory efforts ‘scooping out’ the pressure in the inspiratory phase. Moreover, we observed an increase in the magnitude of the negative esophageal pressure swings for FLs breaths, which aligns with the existing understanding of FL-VD breaths having more pronounced inspiratory efforts. This pressure magnitude was calculated as the minimum esophageal pressure subtracted from the esophageal pressure at the start of the breath.

### FL-VD severity is not conclusively linked to clinical interventions

The FL-VD severity metric is a continuous marker that can inform us about the severity of the dyssynchronous patient-ventilator interaction when the flow provided by the ventilator does not meet patient demands. The state of the patient, such as the severity of illness, and clinical interventions, such as sedation or neuromuscular blockade, influence respiratory drive and the presence of FL-VD. To qualitatively explore this association, we selected three patients out of thirteen and reviewed their treatment history during the analyzed MV period. For each selected patient, the FL-VD severity metric (Fig. 6b-d) and the value *A*_*p*1_ parameter (Fig. 6e-g) are shown in the temporal order as they appeared in the recorded dataset. The frequency of each phenotype of breath averaged for every 30 min time window is shown in Fig. 6h-j.

Patient 104 (Fig. 6b) was ventilated due to pneumonia and a significant increase in the FLs breaths was observed at t=∼18 hour and then again at t=∼24 hour from the time of intubation. The patient was acutely hypotensive during the recording window and there was a general deterioration throughout the recording period attributed to a worsening of shock due to pneumonia. The ventilator was changed from a pressure-controlled volume-targeted mode (APVCMV) at t=14 hours to a patient-triggered pressure support and reverted to APVCMV three hours after the end of the recording window, so it is possible that the ventilator settings are responsible for the period of dyssynchrony. Patient 108 (Fig. 6c) was admitted with an ischemic stroke, then had hemorrhagic conversion 4 days prior to the recording window. Two hours and twenty minutes before recording started, the IV administration of fentanyl (50mcg/hr) and midazolam 0.5mg/h) was stopped and then 15 minutes later, midazolam was restarted at 1mg/hr. Otherwise, vital signs were stable, the critical care observation pain tool (CPOT) score was 0, and ventilator settings were constant. These events do not align with the FL-VD surges at hours 15 and 25 (Fig. 6c). Patient ID 114 (Fig. 6d) was intubated one day prior to the recording window due to pneumonia and empyema. Ventilator settings and sedation were stable prior to and throughout the recording period and thus did not provide any indication of the cause of the period of the severe FL-VD observed in the first ∼4 hours of the recording or the reduction in FL-VD in the remainder of the analysis window. Body temperature was elevated in the period spanning two hours prior to recording through 5 hours into the recording, which overlaps the period of FL-VD.

## DISCUSSION

In this prospective cohort study of mechanically ventilated patients with or at risk for ARDS, we developed a hypothesis-driven mathematical model that targets specific features in the pressure waveform signal associated with FL-VD. An efficient data assimilation (DA)-based pipeline was developed to estimate the feature-specific parameters using a large ventilator waveform dataset. Using the estimated parameters, we successfully developed a formulation to phenotype FL-VD breaths based on their severity with respect to NL breaths. We also analyzed the association between ventilator parameters, healthcare interventions, and the occurrence of FL-VD breaths using the VD severity metric. Critical care physicians often use the ventilator waveform signals to infer the lung condition by visualizing the waveforms and using prior experience to assess the lung conditions and identify different VD types. We tried to systematically mathematize this knowledge by examining the effect of model parameters on the respective pressure and volume waveforms and how those changes associate with FL-VD breaths.

Most previous modeling efforts have not addressed VD and the patient-ventilator interaction directly. Among physiological and non-physiological lung-ventilator models, our methodology’s primary contribution is that it can be used with ventilator waveform data with high variability using the same underlying model formulation as it is driven by the hypothesized patient-ventilator interactions and physiological features. The model quantifies, verifies, and explains both lung state and dyssynchrony relative to the hypothesis that drove the deviation from normal breathing and functions in cases where simple models, e.g., the linear single compartment, lack the flexibility to represent the underlying processes. In this way, our model could act as a bridge between the standard models, which are currently unable to represent clinically observed pathologies such as VD and observed clinical ventilator data.

The temporal pattern of dyssynchronous breaths suggests that discrete events may explain the onset or resolution of dyssynchronous phases. However, a qualitative analysis of the electronic medical record (EMR) did not reveal any striking associations between clinical practice and FL-VD incidence. This is likely due to the fact that the EMR is not a collection of clinical trials or controlled experiments, and the recorded information is subjected to individual variability and unmeasurable confounders. Additionally, temporal resolution in the EMR also limits the determination of the etiology of FL-VD incidence, e.g., sedation scores were recorded every 4 hours. In other words, the EMR does not have enough information to determine which interventions might be better or worse for a patient during VD breaths and such an analysis will require a much larger dataset. But, our model lays the groundwork for future clinical trials that will have the right information to improve ventilator management.

In clinical settings, there are limited lung dysfunction and injury markers, such as compliance, P:F ratio, driving pressure, etc. [47, 48]. Additionally, we have limited methods to identify different types of VD, their frequency and their severity. Therefore, a correlation between the level of underlying lung injury and VD incidence is not well understood [17]. This is further complicated by the interactions between lung injury, ventilator settings, and healthcare processes. This absence of high-resolution and sensitive lung injury markers, and temporal worsening or resolution of lung injury independent of VD, hinders the identification of injury caused by FL-VD. Furthermore, there is an expected latency between FL-VD incidence and injury reflected in, e.g., compliance and the data available for analysis does not encompass the entire ICU stay. Finally, our data were collected for 48 hours after intubation, and do not capture the entire mechanical ventilation course. Accordingly, we did not seek to link the incidence and severity of VD to clinical outcomes. Nevertheless, to identify the causes or effects of VD, first we need to have a machinery that delineates breath into parametric form, which can then be used to demarcate them depending on the severity level. This paper formulates, demonstrates, and validates one approach to solve this problem for FL-VD. The work presented here would allow research into the causes of VD, injury resulting from VD, and the influence of VD on patient outcomes.

The association between different types of VD and ventilator settings is also not well documented. This work showed that the FL-VD severity metric can be used to understand how severe FL breaths affect key features in the pressure and volume waveform signals, which act as a marker to identify FL-VD breaths. We then showed what other model parameters (waveform features) are associated with FL-VD breaths and determined if any specific ventilator setting is highly correlated with severe FL-VD breaths. Therefore, VD-DLV model-defined VD severity metric provides a pathway toward developing methodological machinery for determining VD-related causes and effects. The work proposed here is meant to be a foundation for identifying and quantifying different types of VD. This, in turn, could guide adjusting the ventilator and other clinical practices (e.g., sedation) to reduce VD.

We have demonstrated that the VD-DLV model can accurately represent FL-VD breaths and those fits are used to develop a severity metric that acts as a continuous marker to quantify the severity of FL breaths. Future studies will develop and apply the same methodology to other VD types. Besides FL-VD, at least six other types of VD have been identified in the literature [5, 14]. In the follow-up work, we plan to extend the VD-DLV model to identify and quantify different VD types. While the presented model is developed to be able to target most of the other VD types, we can add new VD-specific sub-models that will act as an estimable term that alters the existing pressure model. For example, reverse and double-triggering VD induces extremely heterogeneous responses in the waveform signals, so minor model expansion might be required [15]. However, modeling other types of VD may necessitate including both pressure and volume parameters in the VD severity calculation as some VD types produce almost similar changes in the pressure signal, such as reverse and double-triggering VD breaths. This will require relevant modification in the estimation methodology to achieve solutions that can converge without by-hand tuning.

We acknowledge several important limitations to this work:1) We did not identify FL-VD breaths in the presence of other VD types. For example, in the presence of delayed cycling VD breaths, the model might not be able to identify FL-VD if we use only the pressure signal model alone [5]. This is not a particular limitation of the model but rather a limitation of this work. The work presented here is meant to be a first step towards developing complex machinery that can simultaneously recognize multiple types of VD breaths. 2) We sampled limited breaths from the continuous ventilator data for optimum estimation. While this was necessary to analyze many patients, time-sensitive information might be missed, given that 100 breaths were sampled out of ∼600 breaths per 30 min. We will investigate further enhancing the computation capabilities and optimizing the estimation methodology to analyze a greater fraction of breaths. 3) The phenotypic bounds of the VD severity metric that determine the severity of FL-VD breaths are for qualitative purposes only. Currently, no standard quantitative method exists to assess the severity of any VD breaths. However, differing opinions will likely exist among experts. Collecting and synthesizing such information will require different future studies. 4) This is a small, single-center study describing results from one specific type of ventilator (Hamilton G5), operated in a pressure-controlled volume-targeted mode, collected for 48 hours after intubation. Additional data verification is required for the model’s broader applicability across different ventilators and ventilation modes. Therefore, conclusions regarding the clinical significance of VD cannot be made without further study.

## CONCLUSION

Using a hypothesis-driven VD-deformed lung ventilator model, we present a novel approach to identify and quantify the severity of flow limited-VD (FL-VD) breaths with respect to NL breaths. This mathematical model uses clinical and physiologic knowledge to define the allowed pressure and volume diversity and accurately reproduces ventilator pressure and volume waveform data of 13 ICU patients. The estimation methodology tracks deformations in pressure signals corresponding to FL-VD breaths so that the VD-DLV model accurately reproduces the characteristic features of flow-limited dyssynchrony. We quantified waveform features at the resolution of the single breath to determine how those features vary over time for different breaths, creating a by-breath parametric representation. We then used those parameters to define the FL-VD severity metric and phenotype FL-VD breaths by severity. The consistency of the phenotypes was verified against an ML-based approach and the rank-order correlation with ventilator parameters was analyzed. A pathophysiological relation between the VD severity metric and healthcare interventions was also explored. In essence, we compute hypothesis-driven breath phenotypes that may potentially be used to run trials on outcomes related to the frequency and strength of certain deviations that correspond to types of VD and hence have direct application to clinical practice and afford meaningful knowledge extraction from clinically collected data.

## GRANTS

This work was supported by National Institutes of Health R01s LM012734 “Mechanistic machine learning,” LM006910 “Discovering and applying knowledge in clinical databases,” HL151630 “Predicting and Preventing Ventilator-Induced Lung Injury” along with R00 HL128944, and K24 HL069223.

## AUTHOR CONTRIBUTION

D.K.A., B.J.S., and D.J.A. conception and design of research; D.K.A and D.J.A. developed the mathematical model, D.K.A., B.J.S., and P.D.S. designed and conducted experiments; D.K.A., B.J.S., and D.J.A. analyzed data and evaluated the model; P.D.S. collected the data; D.K.A., B.J.S., P.D.S., and D.J.A. interpreted results; D.K.A. prepared figures; D.K.A. wrote the original draft of the manuscript; D.K.A., B.J.S., P.D.S., G.H., and D.J.A. edited and revised the manuscript; D.K.A., B.J.S., P.D.S., G.H., and D.J.A. approved the final version of the manuscript.

## Data Availability

All data produced in the present study are available upon reasonable request to the authors.

## REFERENCES

1. Yoshida, T., et al., Fifty years of research in ARDS. Spontaneous breathing during mechanical ventilation. Risks, mechanisms, and management. American journal of respiratory and critical care medicine, 2017. 195(8): p. 985–992.

2. Force, A.D.T., et al., Acute respiratory distress syndrome. Jama, 2012. 307(23): p. 2526–2533.

3. Amato, M.B., et al., Driving pressure and survival in the acute respiratory distress syndrome. New England Journal of Medicine, 2015. 372(8): p. 747–755.

4. Sottile, P.D., D. Albers, and M.M. Moss, Neuromuscular blockade is associated with the attenuation of biomarkers of epithelial and endothelial injury in patients with moderate-to-severe acute respiratory distress syndrome. Critical Care, 2018. 22(1): p. 63.

5. Sottile, P.D., et al., Ventilator dyssynchrony–Detection, pathophysiology, and clinical relevance: A Narrative review. Annals of Thoracic Medicine, 2020. 15(4): p. 190.

6. Gilstrap, D. and N. MacIntyre, Patient–ventilator interactions. Implications for clinical management. American journal of respiratory and critical care medicine, 2013. 188(9): p. 1058–1068.

7. Blanch, L., et al., Asynchronies during mechanical ventilation are associated with mortality. Intensive care medicine, 2015. 41(4): p. 633–641.

8. Aoyama, H., et al., Association of driving pressure with mortality among ventilated patients with acute respiratory distress syndrome: a systematic review and meta-analysis. Critical care medicine, 2018. 46(2): p. 300–306.

9. Bates, J.H. and B.J. Smith, Ventilator-induced lung injury and lung mechanics. Annals of translational medicine, 2018. 6(19).

10. Slutsky, A.S. and V.M. Ranieri, Ventilator-induced lung injury. New England Journal of Medicine, 2013. 369(22): p. 2126–2136.

11. Bellani, G., et al., Epidemiology, patterns of care, and mortality for patients with acute respiratory distress syndrome in intensive care units in 50 countries. Jama, 2016. 315(8): p. 788–800.

12. Corona, T.M. and M. Aumann, Ventilator waveform interpretation in mechanically ventilated small animals. Journal of Veterinary Emergency and Critical Care, 2011. 21(5): p. 496–514.

13. Mellema, M.S., Ventilator waveforms. Topics in companion animal medicine, 2013. 28(3): p. 112–123.

14. Nilsestuen, J.O. and K.D. Hargett, Using ventilator graphics to identify patient-ventilator asynchrony. Respir Care, 2005. 50(2): p. 202–34; discussion 232-4.

15. Daniel, H. and I. Ivan, Identifying patient-ventilator asynchrony using waveform analysis. Palliat Med Care, 2017. 4(4): p. 1–6.

16. Ramirez, I.I., et al., Ability of ICU Health-Care Professionals to Identify Patient-Ventilator Asynchrony Using Waveform Analysis. Respiratory Care, 2017. 62(2): p. 144–149.

17. Mellott, K.G., et al., Patient ventilator asynchrony in critically ill adults: frequency and types. Heart Lung, 2014. 43(3): p. 231–43.

18. Colombo, D., et al., Efficacy of ventilator waveforms observation in detecting patient-ventilator asynchrony. Crit Care Med, 2011. 39(11): p. 2452–7.

19. Georgopoulos, D., G. Prinianakis, and E. Kondili, Bedside waveforms interpretation as a tool to identify patient-ventilator asynchronies. Intensive Care Medicine, 2006. 32(1): p. 34–47.

20. Liao, K.M., C.Y. Ou, and C.W. Chen, Classifying different types of double triggering based on airway pressure and flow deflection in mechanically ventilated patients. Respir Care, 2011. 56(4): p. 460–6.

21. Zhang, L., et al., Detection of patient-ventilator asynchrony from mechanical ventilation waveforms using a two-layer long short-term memory neural network. Comput Biol Med, 2020. 120: p. 103721.

22. Blanch, L., et al., Validation of the Better Care system to detect ineffective efforts during expiration in mechanically ventilated patients: a pilot study (vol 38, pg 772>, 2012). Intensive Care Medicine, 2013. 39(2): p. 341-341.

23. Rolland-Debord, C., et al., Prevalence and Prognosis Impact of Patient-Ventilator Asynchrony in Early Phase of Weaning according to Two Detection Methods. Anesthesiology, 2017. 127(6): p. 989–997.

24. Pan, Q., et al., Identifying Patient-Ventilator Asynchrony on a Small Dataset Using Image-Based Transfer Learning. Sensors (Basel), 2021. 21(12).

25. Mellenthin, M.M., et al., Using injury cost functions from a predictive single-compartment model to assess the severity of mechanical ventilator-induced lung injuries. Journal of Applied Physiology, 2019. 127(1): p. 58–70.

26. Hamlington, K.L., et al., Predicting ventilator-induced lung injury using a lung injury cost function. Journal of Applied Physiology, 2016. 121(1): p. 106–114.

27. van Diepen, A., et al., A model-based approach to generating annotated pressure support waveforms. Journal of Clinical Monitoring and Computing, 2022. 36(6): p. 1739–1752.

28. Athanasiades, A., et al., Energy analysis of a nonlinear model of the normal human lung. Journal of Biological Systems, 2000. 8(02): p. 115–139.

29. de Wit, M., et al., Observational study of patient-ventilator asynchrony and relationship to sedation level. Journal of Critical Care, 2009. 24(1): p. 74–80.

30. Agrawal, D.K., et al., A Damaged-Informed Lung Ventilator Model for Ventilator Waveforms. Frontiers in Physiology, 2021. 12: p. 724046.

31. Antonogiannaki, E.M., D. Georgopoulos, and E. Akoumianaki, Patient-Ventilator Dyssynchrony. Korean J Crit Care Med, 2017. 32(4): p. 307–322.

32. Sottile, P.D., et al., The Association Between Ventilator Dyssynchrony, Delivered Tidal Volume, and Sedation Using a Novel Automated Ventilator Dyssynchrony Detection Algorithm. Critical Care Medicine, 2018. 46(2): p. E151–E157.

33. Pohlman, M.C., et al., Excessive tidal volume from breath stacking during lung-protective ventilation for acute lung injury. Critical care medicine, 2008. 36(11): p. 3019–3023.

34. Grasso, S., et al., Airway pressure-time curve profile (stress index) detects tidal recruitment/hyperinflation in experimental acute lung injury. Crit Care Med, 2004. 32(4): p. 1018–27.

35. Sottile, P.D., et al., The association between ventilator dyssynchrony, delivered tidal volume, and sedation using a novel automated ventilator dyssynchrony detection algorithm. Critical care medicine, 2018. 46(2): p. e151.

36. Cavalcanti, A.B., et al., Effect of lung recruitment and titrated positive end-expiratory pressure (PEEP) vs low PEEP on mortality in patients with acute respiratory distress syndrome: a randomized clinical trial. Jama, 2017. 318(14): p. 1335–1345.

37. Baydur, A., et al., A simple method for assessing the validity of the esophageal balloon technique. American Review of Respiratory Disease, 1982. 126(5): p. 788–791.

38. Bertsimas D, T.J., Introduction to Linear Optimization. 1997: Athena Scientific.

39. Nocedal J, a.W.S., Numerical Optimization. 2006: Springer, New York.

40. Nelder, J.A. and R. Mead, A simplex method for function minimization. The computer journal, 1965. 7(4): p. 308–313.

41. Albers, D.J., et al., Ensemble Kalman methods with constraints. Inverse Problems, 2019. 35(9): p. 095007.

42. Smith, R.C., Uncertainty quantification: theory, implementation, and applications. Vol. 12. 2013: Siam.

43. Law, K., A. Stuart, and K. Zygalakis, Data assimilation. Cham, Switzerland: Springer, 2015.

44. Asch, M., M. Bocquet, and M. Nodet, Data assimilation: methods, algorithms, and applications. 2016: SIAM.

45. Albers, D.J., et al., The Parameter Houlihan: a solution to high-throughput identifiability indeterminacy for brutally ill-posed problems. Mathematical biosciences, 2019. 316: p. 108242.

46. MacIntyre, N.R., et al., Patient-ventilator flow dyssynchrony: Flow-limited versus pressure-limited breaths. Critical Care Medicine, 1997. 25(10): p. 1671–1677.

47. Chiumello, D., et al., Airway driving pressure and lung stress in ARDS patients. Crit Care, 2016. 20: p. 276.

48. Smith, B.J., et al., Predicting the response of the injured lung to the mechanical breath profile. Journal of applied physiology, 2015. 118(7): p. 932–940.

