## Supplementary Information for "Quantifiable identification of flow-limited ventilator dyssynchrony with the deformed lung ventilator model"

**Deepak. K. Agrawal[1,6*], Bradford J. Smith[2,3], Peter D. Sottile[4], George Hripcsak[5], and David J. Albers[2,5,6*]**
[1]Centre for Biomedical Engineering, Indian Institute of India Delhi, New Delhi, 110016, India
[2]Department of Bioengineering, University of Colorado Denver | Anschutz Medical Campus, Aurora, CO 80045, USA
[3]Section of Pulmonary and Sleep Medicine, Department of Pediatrics, University of Colorado Denver, Anschutz Medical Campus, Aurora, CO 80045, USA
[4]Division of Pulmonary Sciences and Critical Care Medicine, Department of Medicine, University of Colorado School of Medicine, Aurora, CO 80045, USA
[5]Department of Biomedical Informatics, Columbia University, New York, NY USA
[6]Section of Informatics and Data Science, Department of Pediatrics, School of Medicine, University of Colorado Denver, Anschutz Medical Campus, Aurora, CO 80045, USA

***Correspondence:**
Corresponding Authors


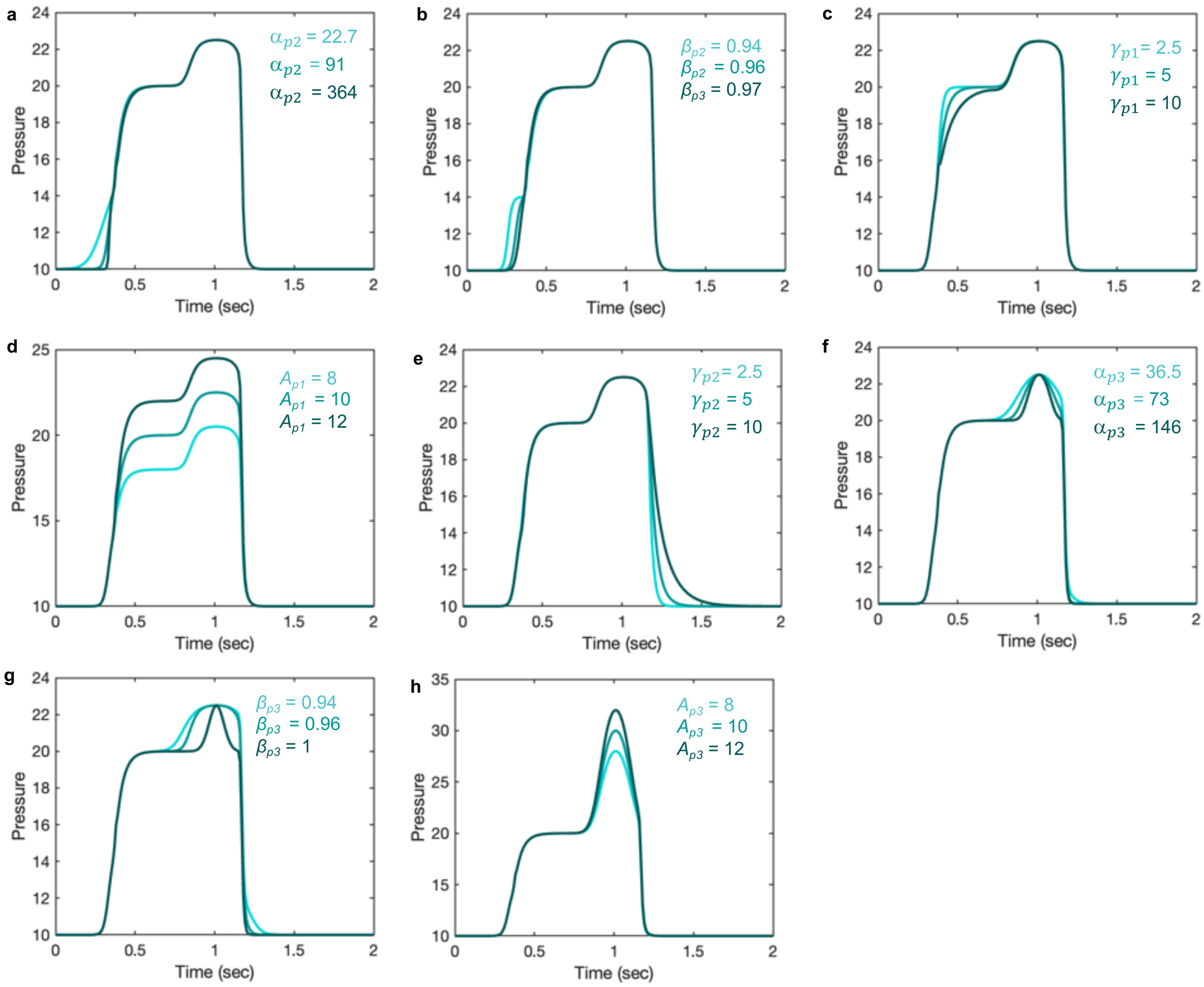


**Figure S1: Demonstrating the pressure model flexibility by altering relevant features in the pressure waveform.** The initial gradient of the pressure signal during inspiration at low volume is controlled by the **(a)** $\alpha_{p2}$ and **(b)** $\beta_{p2}$ parameters. Higher values result in a slower rise. **(c)** The gradient of the rising signal after the inflection point is controlled by the $\gamma_{p1}$ parameter. Higher values result in a slower rise. **(d)** The amplitude of the waveform can be altered using the parameter $A_{p1}$. **(e)** The gradient of the falling signal during expiration can be modified by the $\gamma_{p2}$ parameter. Higher values result in a slower fall. The deformation at the plateau is regulated by the **(f)** $\alpha_{p3}$ ($\beta_{p3}$ = 0.999) and **(g)** $\beta_{p3}$ parameters such that higher values result in a sharper peak. **(i)** The amplitude of this deformation is controlled via $A_{p3}$ parameter. Equations 1-10 were used to simulate the response within each term with parameter values $\theta$ = 0.2, $\alpha_{p1}$= 189, $\beta_{p1}$ = 0.88, $\varphi_{p1}$ = -0.6, $\gamma_{p1}$= 5, $\gamma_{p2}$= 1.52, $A_{p1}$ = 10, $CP$ = 10, $\alpha_{p2}$= 91, $\beta_{p2}$ = 0.97, $\varphi_{p2}$ = -0.9, $A_{p2}$ = 4, $\alpha_{p3}$= 73, $\beta_{p3}$ = 0.98, $\varphi_{p3}$ = -0.3, $A_{p3}$ = 2.5 otherwise. Note that the model variability shown here is independent of the ventilator mode.

### Section I: Constructing VD-deformed volume model

The basic framework of the volume model remains the same as the pressure model, where the deformed volume waveform model is derived from the periodic rectangular waveform signal. These signals are produced by combining the sinusoidal function with the hyperbolic tangent function. These rectangular waveform signals allow us to independently control the rate of inspiration or expiration part of the breath while maintaining the periodicity. The rectangular waveform signal is produced by:

$$f_{vb} = \frac{1}{2}\{\tanh(\alpha_{v1}\,(\sin(2\pi\theta t - \varphi_{v1}) - \beta_{v1}) + 1\} \qquad \text{(S1)}$$

Here, the parameters $\alpha_{v1}$ , $\phi_{v1}$ and $\beta_{v1}$ allow controlling the smoothness, starting point and duty cycle of the rectangular waveform, respectively. The other terms (1/2, +1) are added to generate a rectangular waveform with zero-base value and unit amplitude. The volume model is described as

$$V = f_{v11} + f_{v21} + f_{v32} \qquad \text{(S2)}$$

where the $f_{v11}$ and $f_{v21}$ components produce the inspiration (feature A) and the expiration (feature B) part of the volume signal (Fig. 2b), respectively while the $f_{v32}$ component produces an additional bump that could be used to reproduce dyssynchrony-related features in the volume signal, such as observed in double-triggered breaths (Fig. S2b).

$$f_{v11}(i+1) = \left\{\frac{1}{\gamma_{v1}}\, f_{vb}(i+1) + \left(1 - \frac{1}{\gamma_{v1}}\right) f_{v11}(i)\right\}; \quad i = 1{:}\,n,$$
$$f_{v11} = A_{v1}[f_{v11}(1)\, f_{v11}(2) \ldots\, f_{v11}(i) \ldots f_{v11}(n)]\, \frac{f_{vb}}{\max[f_{v11}]} \qquad \text{(S3)}$$

$$f_{v21}(i+1) = \left\{\frac{1}{\gamma_{v2}}\, f_{vb}(i+1) + \left(1 - \frac{1}{\gamma_{v2}}\right) f_{v21}(i)\right\}; \quad i = 1{:}\,n,$$
$$f_{v21} = A_{v1}[f_{v21}(1)\, f_{v21}(2) \ldots\, f_{v21}(i) \ldots f_{v12}(n)]\, \frac{f_{vb}}{\max[f_{v21}]} \qquad \text{(S4)}$$

$$f_{v32} = A_{v2}\frac{f_{v31}}{\max[f_{v31}]} \qquad \text{(S5)}$$
$$f_{v31} = \frac{1}{2}\{\tanh(\alpha_{v2}\,(\sin(2\pi\theta t - \varphi_{v2}) - \beta_{v2})) + 1\} \qquad \text{(S6)}$$

A short description of how the model parameters contribute to the model is provided in Table S1, and how each of the model components contributes to the deformed model is shown in Fig. S2. Figure S3 shows how some of the important model parameters such as $\gamma_{v1}$, $\gamma_{v2}$, and $A_{v1}$ impact the volume waveform in a targeted way and have interpretable pathophysiology with hypothesized lung conditions that clinicians often use.

**Table S1:** Interpreting VD-deformed lung ventilator (DLV) volume model parameters. The parameters that are correlated with known measures of lung physiology are in bold.

| Parameter | Model relevance | Physiological relevance (with increased values) |
|---|---|---|
| $\theta$ | Number of breaths/s. Higher values result in a higher number of breaths/s. | - |
| $\alpha_{v1}$ | Smoothness of the square waveform ($f_{vb}$). Higher values result in a sharper transition. | - |
| $\beta_{v1}$ | I:E ratio. Higher values result in a smaller duration of inspiration. | - |
| $\varphi_{v1}$ | Starting of the inspiration point. Higher value results in a more delayed response. | - |
| $\gamma_{v1}$ | Gradient of the rising signal. Higher values result in a slower rising signal. | Lower compliance and/or higher resistance. |
| $\gamma_{v2}$ | Gradient of the falling signal. Higher values result in a slower falling signal | Higher (slower) expiratory time constant. |
| $\alpha_{v2}$, $\beta_{v2}$, $\varphi_{v2}$ | Gradient, width, and position of the bump produced by the $f_{v32}$ component | Associated with ventilator desynchrony. |
| $A_{v1}$ | Peak amplitude | Higher overall compliance. |
| $A_{v2}$ | Amplitude of the bump produced by the $f_{v32}$ component | |

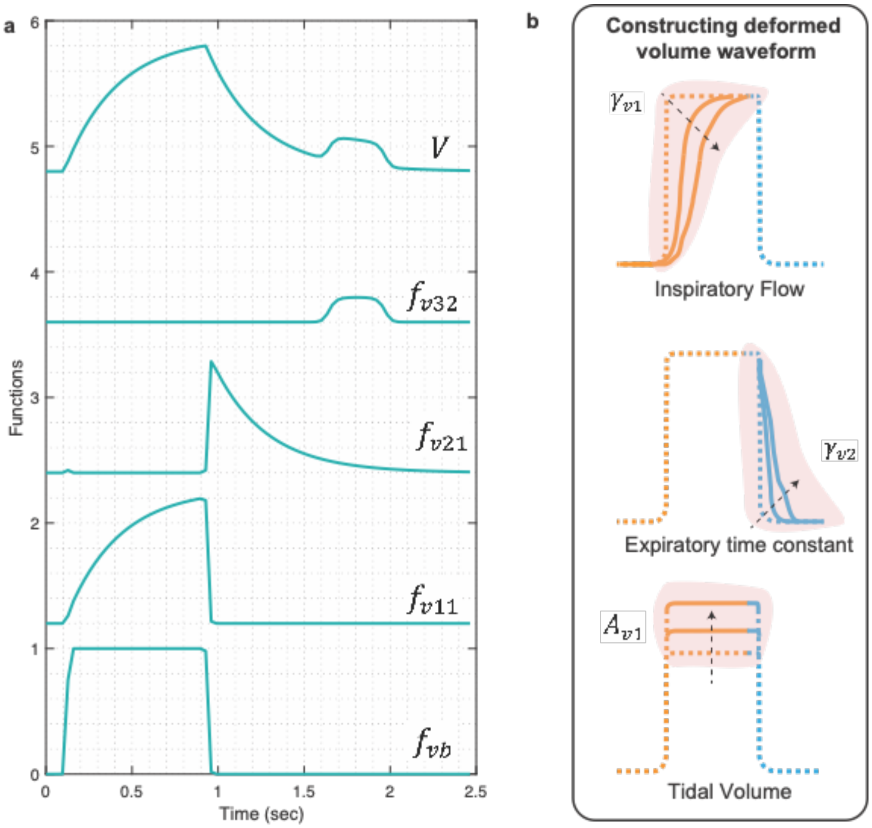


**Figure S2: (a)** Simulated response of various terms that make up the VD-DLV volume model. Equations S1-S6 were used to simulate the response within each term with parameter values $\theta$ = 0.2, $\alpha_{v1}$ = 200, $\boldsymbol{\beta_{v1}}$ = 0.87, $\boldsymbol{\varphi_{v1}}$ = -0.9, $\boldsymbol{\gamma_{v1}}$= 10, $\gamma_{v2}$= 20, $\boldsymbol{A_{v1}}$ = 1, $\alpha_{v2}$ = 100, $\boldsymbol{\beta_{v2}}$ = 0.98,$\varphi_{v2}$ = 0.7, $\boldsymbol{A_{v1}}$ = 0.2, otherwise. Note that the model variability shown here is independent of the ventilator mode. **(b)** In the volume signal, the gradient of the rising (feature A) and falling (feature B) signals can be altered using the $\boldsymbol{\gamma_{v1}}$ and $\gamma_{v2}$ parameters, respectively. Increased values of these parameters increase the transient time for the signal to reach the same volume level. The amplitude of the waveform can be altered using the parameter $\boldsymbol{A_{v1}}$.

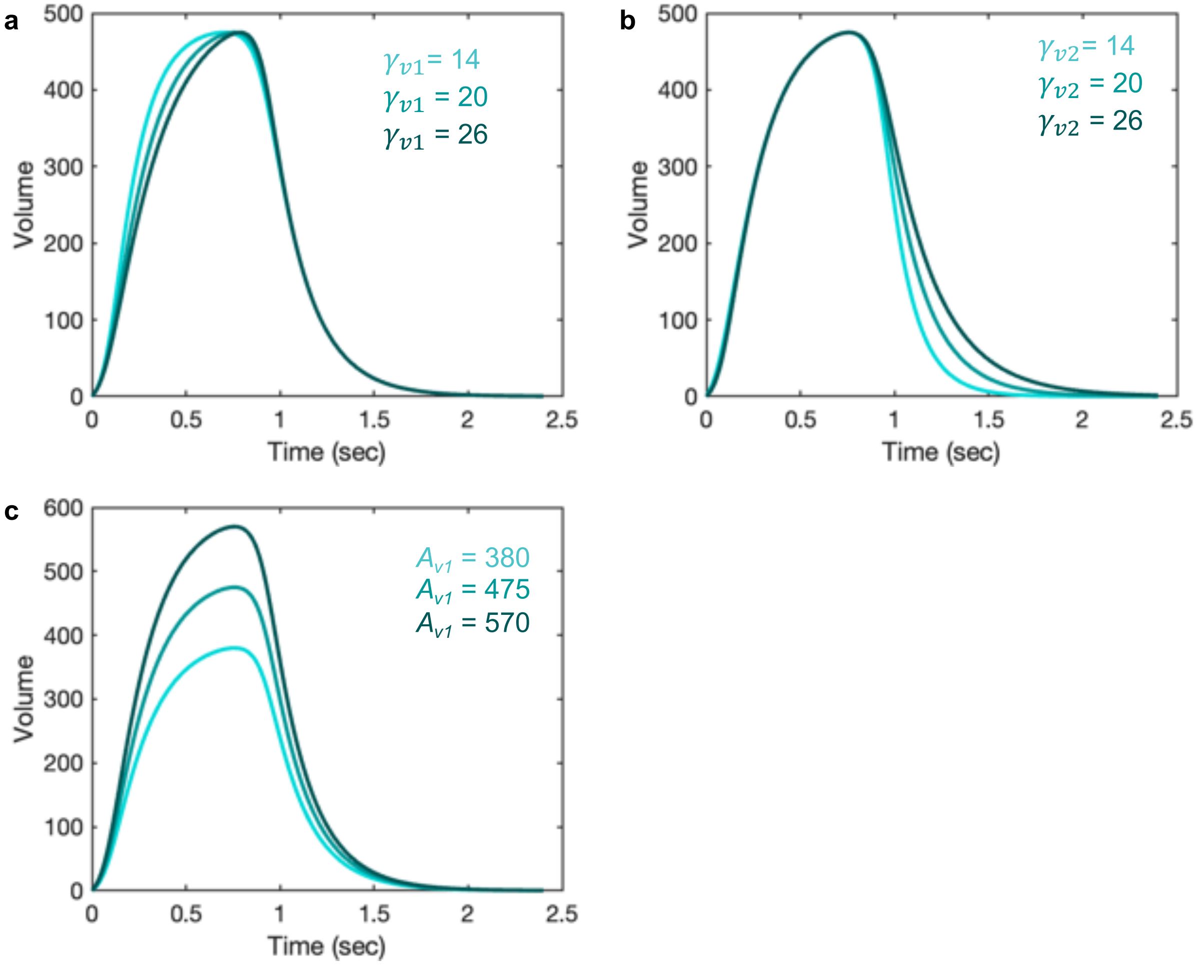


**Figure S3: Demonstrating the volume model flexibility by altering relevant features in the pressure waveform.** In volume signal, parameters $\boldsymbol{\gamma_{v1}}$, $\boldsymbol{\gamma_{v2}}$, and $\boldsymbol{A_{v1}}$ might have physiological meaning when the volume signal is not controlled via a ventilator. **(a)** The parameter, $\boldsymbol{\gamma_{v1}}$ might be inversely correlated with lung compliance while **(b)** the parameter $\boldsymbol{\gamma_{v2}}$, is directly proportional to the expiratory time constant, which is the product of resistance and compliance. **(c)** The parameter $\boldsymbol{A_{v1}}$ is generally referred to as tidal volume and has a direct correlation with compliance. Equations S1-S6 were used to simulate the response within each term with parameter values $\theta$ = 0.2, $\alpha_{v1}$ = 20, $\boldsymbol{\beta_{v1}}$ = 0.87, $\boldsymbol{\varphi_{v1}}$ = -0.95, $\boldsymbol{\gamma_{v1}}$= 20, $\gamma_{v2}$= 20, $\boldsymbol{A_{v1}}$ = 10, $\alpha_{v2}$ = 247, $\beta_{v2}$ = 0.9, $\varphi_{v2}$ = -0.96, $\boldsymbol{A_{v2}}$ = 0, otherwise. Note that the model variability shown here is independent of the ventilator mode.

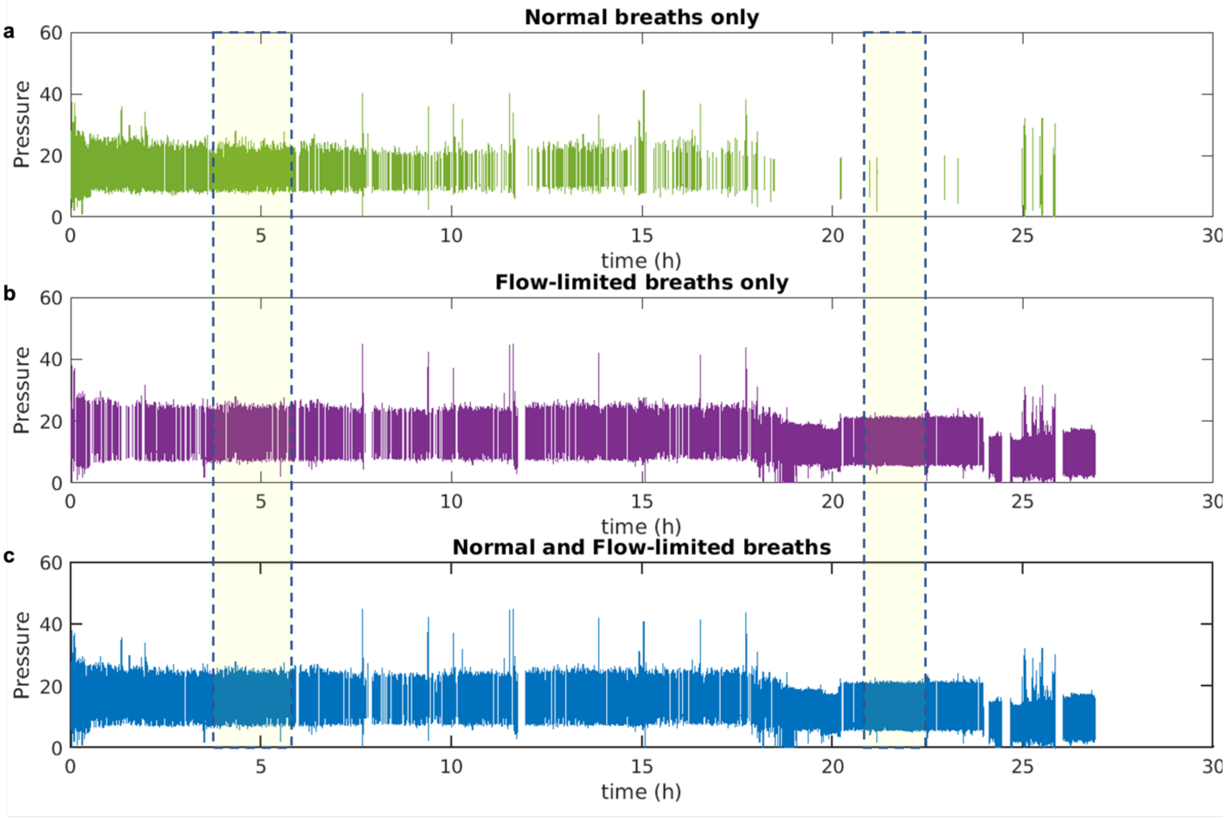


**Figure S4: Recorded breath history of patient ID 104 with normal (NL) and flow-limited (FL) breaths only. (a)** Only normal breaths; **(b)** only flow-limited breaths and **(c)** normal and flow-limited breaths combined are shown as they appeared in the recorded datasets, starting with the first breath at t=0. In the highlighted regions, there are several 30 min sampled time windows where the total number of normal (NL) and flow-limited (FL) breaths combined were more than 100 per 30 min interval. From this filtered dataset, 100 breaths per 30 min sampled window were selected using the RAND function in MATLAB. The process was repeated for the entire recorded time of the dataset.

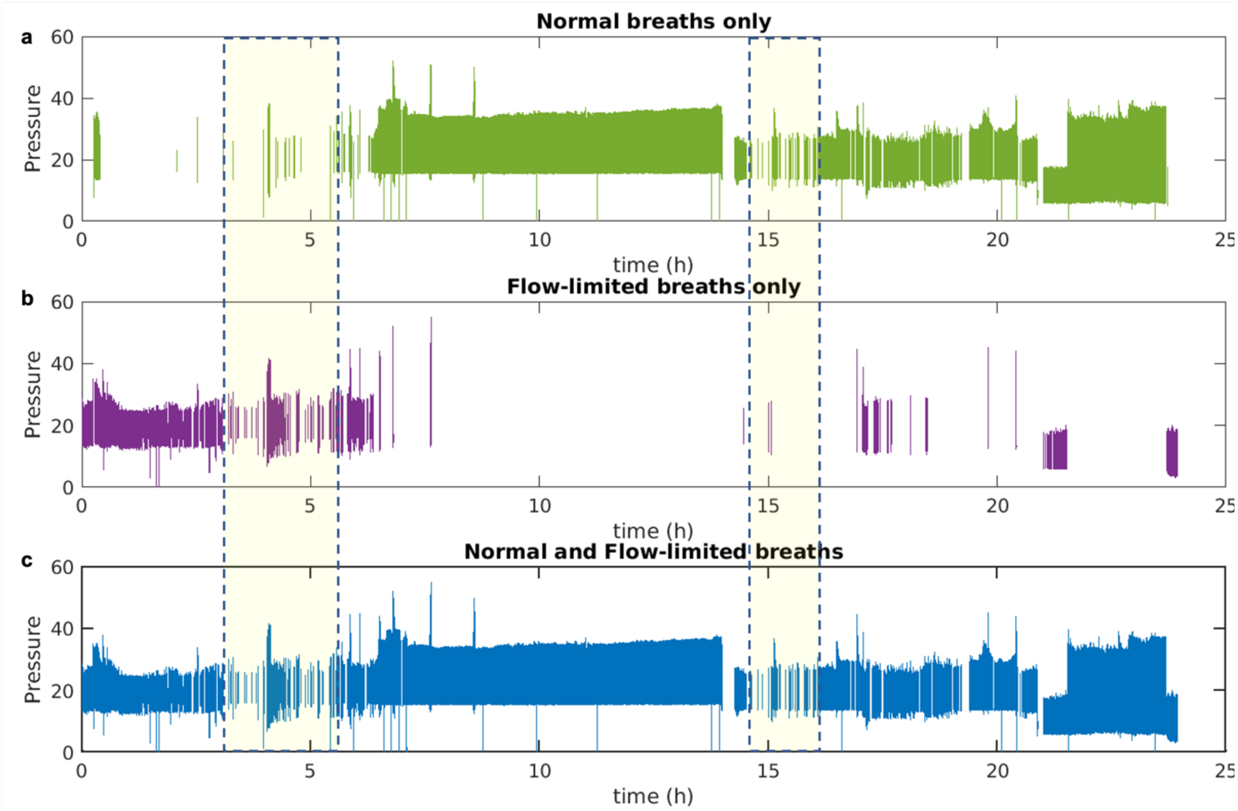


**Figure S5: Recorded breath history of patient ID 114 with normal (NL) and flow-limited (FL) breaths only. (a)** Only normal breaths; **(b)** only flow-limited breaths and **(c)** normal and flow-limited breaths combined are shown as they appeared in the recorded datasets, starting with the first breath at t=0. In the highlighted regions, there are several 30 min sampled time windows where the total number of NL and FL breaths combined were less than 100 per 30 min interval. From this filtered dataset, the maximum number of available breaths was selected, which ranged from 1 to 100, using the RAND function in MATLAB. The process was repeated for the entire recorded time of the dataset.

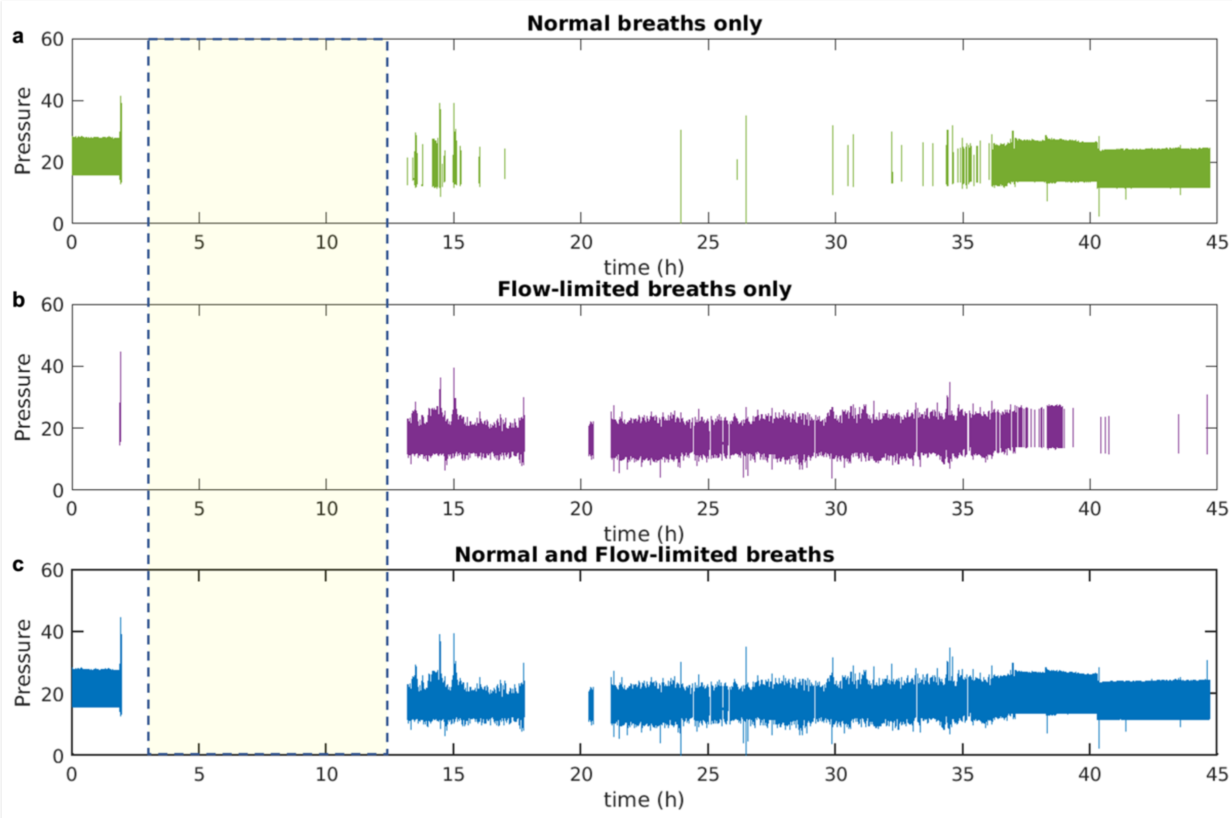


**Figure S6: Recorded breath history of patient ID 129 with normal (NL) and flow-limited (FL) breaths only. (a)** Only normal breaths; **(b)** only flow-limited breaths and **(c)** normal and flow-limited breaths combined are shown as they appeared in the recorded datasets, starting with the first breath at t=0. In the highlighted regions, there are several 30 min sampled time windows where no NL and FL breaths were present. Therefore, no breaths were selected for these regions. The process was repeated for the entire recorded time of the dataset.

**Table S2:** Adjusting the constraints of the $\varphi_{p3}$ parameter in different *Runs* to ensure the correct placement of the $f_{p32}$ component.

| **Run Number** | **Minimum value** | **Maximum value** |
|---|---|---|
| Run 2 | -0.8 | 1.7 |
| Run 3 | -0.8 | -0.3 |
| Run 4 | -0.8 | -0.5 |
| Run 5 | -0.8 | -0.65 |

**Table S3:** Estimated model parameters obtained from the optimization scheme for the results shown in Figure 3.

| Parameters | Case 1 - Run 4 (Fig. 3b) | Case 2 - Run 3 (Fig. 3c) | Case 3 - Run 2 (Fig. 3d) |
|---|---|---|---|
| $\theta$ | 0.2 | 0.2 | 0.2 |
| $\alpha_{v1}$ | 20.0 | 20.0 | 20.0 |
| $\beta_{v1}$ | 0.9 | 0.9 | 0.8 |
| $\varphi_{v1}$ | -0.9 | -0.9 | -0.8 |
| $\gamma_{v1}$ | 9.5 | 5.1 | 11.3 |
| $\gamma_{v2}$ | 7.8 | 18.6 | 20.0 |
| $A_{v1}$ | 445.4 | 408.5 | 446.9 |
| $\alpha_{v2}$ | - | - | - |
| $\beta_{v2}$ | - | - | - |
| $\varphi_{v2}$ | - | - | - |
| $A_{v2}$ | - | - | - |
| $\alpha_{p1}$ | 166.9 | 196.8 | 123.6 |
| $\beta_{p1}$ | 0.9 | 0.9 | 0.8 |
| $\varphi_{p1}$ | -1.0 | -1.0 | -0.7 |
| $\alpha_{p2}$ | 97.7 | 98.3 | 89.8 |
| $\beta_{p2}$ | 1.0 | 1.0 | 1.0 |
| $\varphi_{p2}$ | -1.3 | -1.2 | -1.2 |
| $\gamma_{p1}$ | 2.6 | 2.9 | 4.3 |
| $\gamma_{p2}$ | 2.8 | 1.3 | 3.4 |
| $A_{p1}$ | 20.7 | 14.7 | 3.5 |
| $A_{p2}$ | 1.2 | 1.7 | 1.7 |
| $CP$ | 10.9 | 9.3 | 8.2 |
| $\alpha_{p3}$ | - | 27.6 | 35.9 |
| $\beta_{p3}$ | - | 1.0 | 1.0 |
| $\varphi_{p3}$ | - | -0.8 | -0.3 |
| $A_{p3}$ | - | 3.4 | 5.2 |

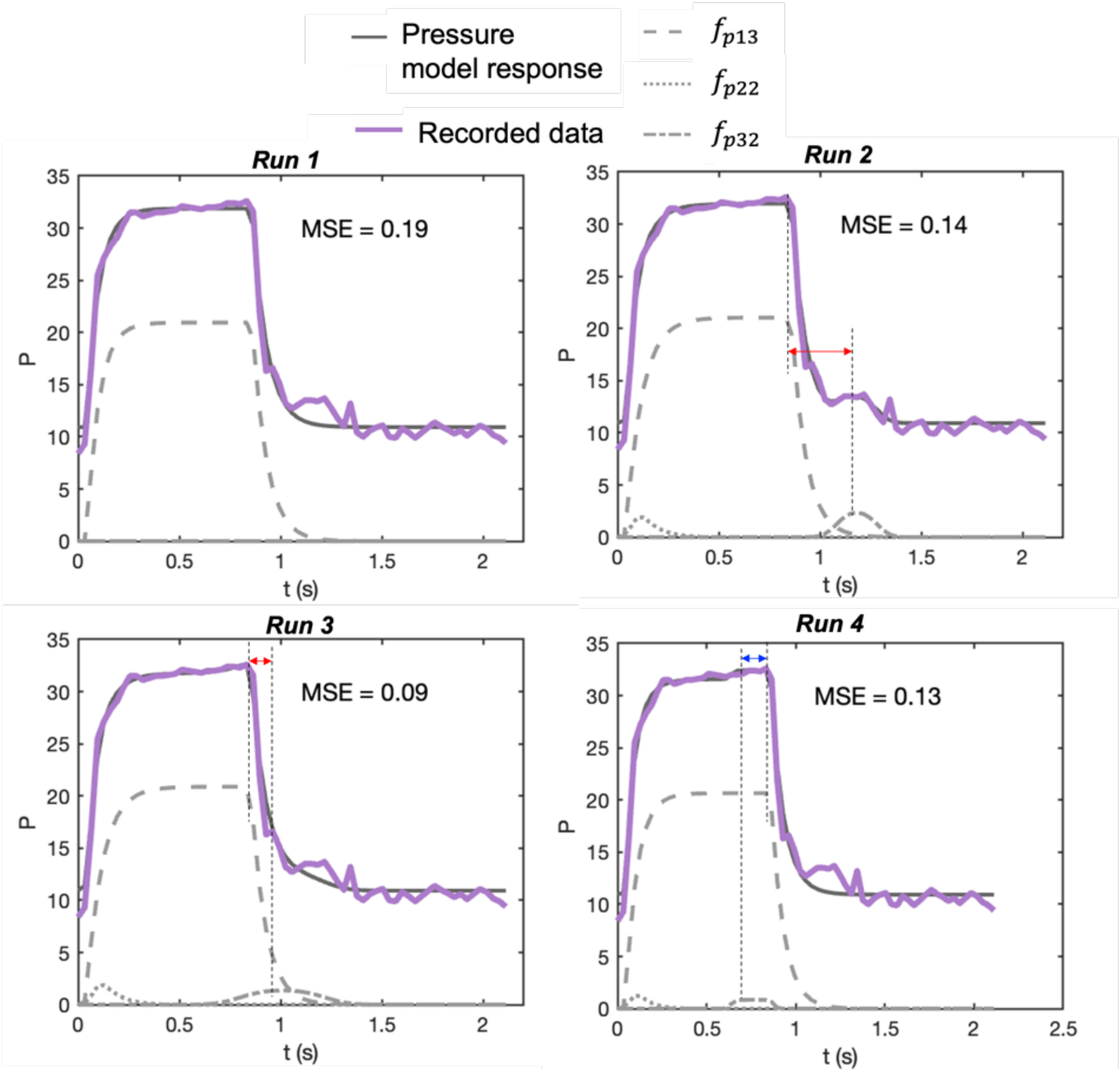


**Figure S7:** The pressure waveforms of the breath representing Case 1 in Fig. 3. The model's response is shown in sold-grey lines, while the recorded data is shown in solid-colored lines. In *Run 1*, only the parameters associated with the baseline model were estimated, while from *Run 2* onwards, all the model parameters were estimated. The pressure signal baseline model comprises the $f_{p13}$ component and $CP$, while the deformed model comprises all three components ($f_{p13}$, $f_{p22}$, and $f_{p32}$) and $CP$. From *Run 3* onwards, only the constraints of the $\varphi_{p3}$ parameter were modified (Table S2) while keeping the constraints of the rest of the parameters the same. This is to ensure the deformation produced by the $f_{p32}$ component is placed during or at the end of inspiration. The correct placement of the $f_{p32}$ component is accepted only when the difference between the end of the inspiration signal and the maximum of the $f_{p32}$ component response is positive- shown by the blue arrow. Otherwise, a red arrow indicates a negative difference. The estimated parameters for each Run are shown in Table S4.

**Table S4:** Estimated model parameters obtained from the optimization scheme for the breath representing Case 1 in Fig. 3. In *Run 1*, only the baseline model (Equation 3 in the main text) parameters were estimated, while from *Run 2* onwards, all the model parameters were estimated. From *Run 3* onwards, only the constraints of the $\varphi_{p3}$ parameter were modified while keeping the constraints of the rest of the parameters the same.

| **Parameters** | **Case 1 - *Run 1*** | **Case 1 - *Run 2*** | **Case 1 - *Run 3*** | **Case 1 - *Run 4*** |
|---|---|---|---|---|
| $\theta$ | 0.2 | 0.2 | 0.2 | 0.2 |
| $\alpha_{v1}$ | 20.0 | 20.0 | 20.0 | 20.0 |
| $\beta_{v1}$ | 0.9 | 0.9 | 0.9 | 0.9 |
| $\varphi_{v1}$ | -0.8 | -0.9 | -0.9 | -0.9 |
| $\gamma_{v1}$ | 10.0 | 9.5 | 9.5 | 9.5 |
| $\gamma_{v2}$ | 5.0 | 7.8 | 7.8 | 7.8 |
| $A_{v1}$ | 449.4 | 445.4 | 445.4 | 445.4 |
| $\alpha_{v2}$ | 10.0 | 10.0 | 10.0 | 10.0 |
| $\beta_{v2}$ | 0.9 | 0.9 | 0.9 | 0.9 |
| $\varphi_{v2}$ | 2.0 | 2.0 | 2.0 | 2.0 |
| $A_{v2}$ | 200.0 | 200.0 | 200.0 | 200.0 |
| $\alpha_{p1}$ | 192.1 | 159.7 | 116.3 | 166.9 |
| $\beta_{p1}$ | 0.9 | 0.9 | 0.9 | 0.9 |
| $\varphi_{p1}$ | -1.0 | -1.0 | -1.0 | -1.0 |
| $\alpha_{p2}$ | - | 98.7 | 99.4 | 97.7 |
| $\beta_{p2}$ | - | 1.0 | 1.0 | 1.0 |
| $\varphi_{p2}$ | - | -1.3 | -1.3 | -1.3 |
| $\gamma_{p1}$ | 2.4 | 2.9 | 3.0 | 2.6 |
| $\gamma_{p2}$ | 2.8 | 2.8 | 2.8 | 2.8 |
| $A_{p1}$ | 20.9 | 21.0 | 20.9 | 20.7 |
| $A_{p2}$ | 0.0 | 1.9 | 1.9 | 1.2 |
| $CP$ | 10.9 | 10.9 | 10.9 | 10.9 |
| $\alpha_{p3}$ | - | 72.0 | 20.3 | 260.4 |
| $\beta_{p3}$ | - | 1.0 | 1.0 | 1.0 |
| $\varphi_{p3}$ | - | **-0.1** | **-0.3** | **-0.6** |
| $A_{p3}$ | - | 2.4 | 1.4 | 0.8 |

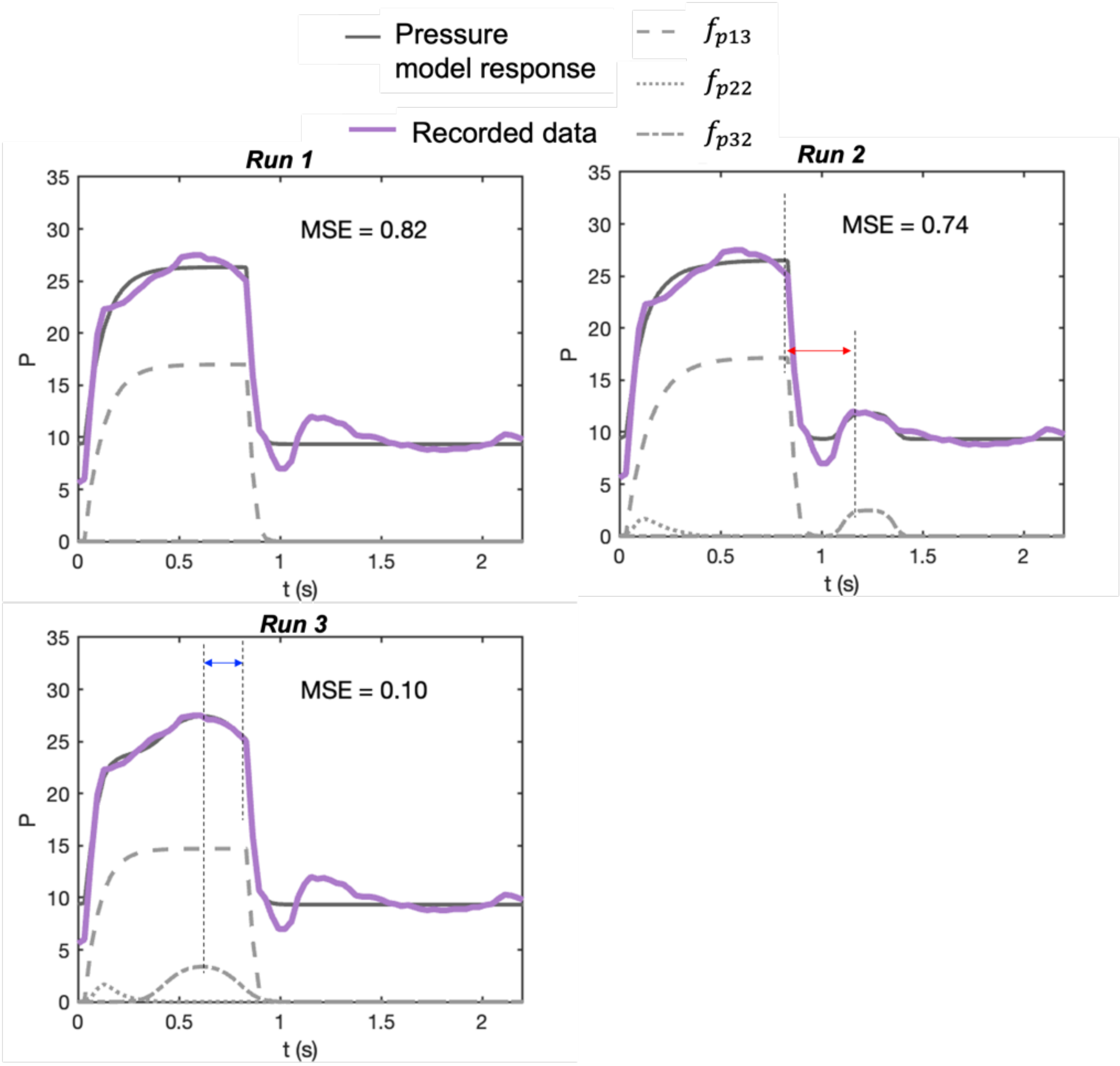


**Figure S8:** The pressure waveforms of the breath representing Case 1 in Fig. 3. The model's response is shown in sold-grey lines, while the recorded data is shown in solid-colored lines. In *Run 1*, only the parameters associated with the baseline model were estimated, while from *Run 2* onwards, all the model parameters were estimated. The pressure signal baseline model comprises the $f_{p13}$ component and $CP$, while the deformed model comprises all three components ($f_{p13}$, $f_{p22}$, and $f_{p32}$) and $CP$. From *Run 3* onwards, only the constraints of the $\varphi_{p3}$ parameter were modified (Table S2) while keeping the constraints of the rest of the parameters the same. This is to ensure the deformation produced by the $f_{p32}$ component is placed during or at the end of inspiration. The correct placement of the $f_{p32}$ component is accepted only when the difference between the end of the inspiration signal and the maximum of the $f_{p32}$ component response is positive- shown by the blue arrow. Otherwise, a red arrow indicates a negative difference. The estimated parameters for each *Run* are shown in Table S5.

**Table S5:** Estimated model parameters obtained from the optimization scheme for the breath representing Case 2 in Figure 3. In *Run 1*, only the baseline model (Equation 3 in the main text) parameters were estimated, while from *Run 2* onwards, all the model parameters were estimated. From *Run 3* onwards, only the constraints of the $\varphi_{p3}$ parameter were modified while keeping the constraints of the rest of the parameters the same.

| **Parameters** | **Case 2 - *Run 1*** | **Case 2 - *Run 2*** | **Case 12- *Run 3*** |
|---|---|---|---|
| $\theta$ | 0.2 | 0.2 | 0.2 |
| $\alpha_{v1}$ | 20.0 | 20.0 | 20.0 |
| $\beta_{v1}$ | 0.9 | 0.9 | 0.9 |
| $\varphi_{v1}$ | -0.8 | -0.9 | -0.9 |
| $\gamma_{v1}$ | 10.0 | 5.1 | 5.1 |
| $\gamma_{v2}$ | 5.0 | 18.6 | 18.6 |
| $A_{v1}$ | 471.1 | 408.5 | 408.5 |
| $\alpha_{v2}$ | 10.0 | 10.0 | 10.0 |
| $\beta_{v2}$ | 0.9 | 0.9 | 0.9 |
| $\varphi_{v2}$ | 2.0 | 2.0 | 2.0 |
| $A_{v2}$ | 200.0 | 200.0 | 200.0 |
| $\alpha_{p1}$ | 196.8 | 190.2 | 196.8 |
| $\beta_{p1}$ | 0.9 | 0.9 | 0.9 |
| $\varphi_{p1}$ | -1.0 | -1.0 | -1.0 |
| $\alpha_{p2}$ | - | 95.9 | 98.3 |
| $\beta_{p2}$ | - | 1.0 | 1.0 |
| $\varphi_{p2}$ | - | -1.3 | -1.2 |
| $\gamma_{p1}$ | 3.3 | 4.1 | 2.9 |
| $\gamma_{p2}$ | 1.3 | 1.3 | 1.3 |
| $A_{p1}$ | 17.0 | 17.1 | 14.7 |
| $A_{p2}$ | 0.0 | 1.7 | 1.7 |
| $CP$ | 9.3 | 9.3 | 9.3 |
| $\alpha_{p3}$ | - | 126.7 | 27.6 |
| $\beta_{p3}$ | - | 1.0 | 1.0 |
| $\varphi_{p3}$ | - | **0.0** | **-0.8** |
| $A_{p3}$ | - | 2.5 | 3.4 |

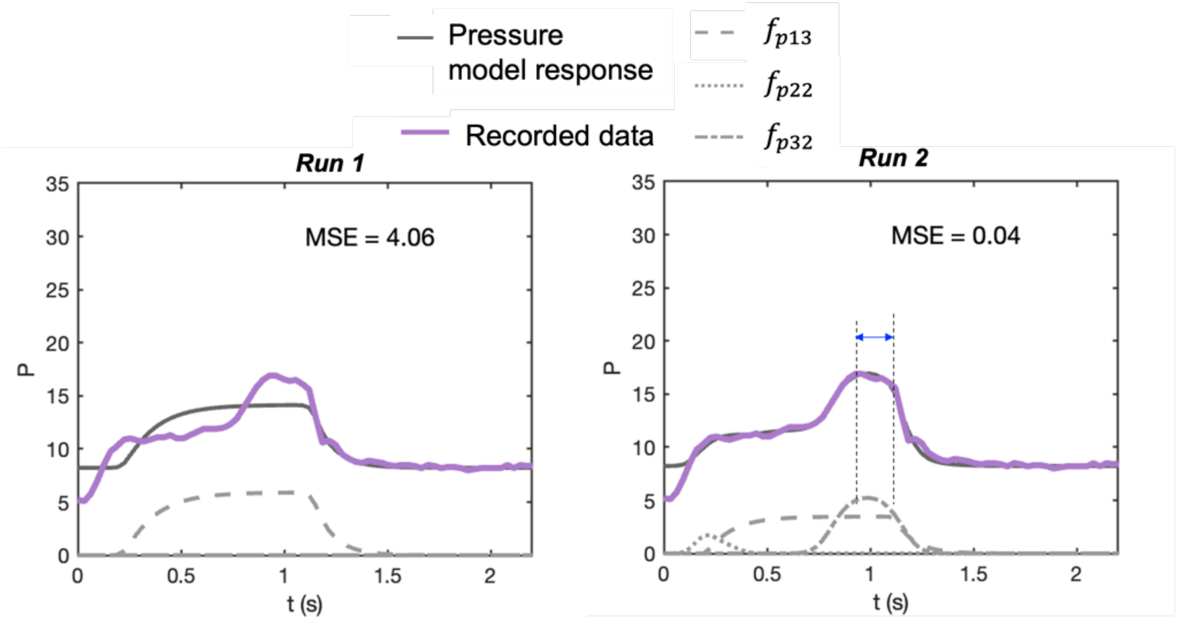


**Figure S9:** The pressure waveforms of the breath representing Case 1 in Fig. 3. The model's response is shown in sold-grey lines, while the recorded data is shown in solid-colored lines. In *Run 1*, only the parameters associated with the baseline model were estimated, while from *Run 2* onwards, all the model parameters were estimated. The pressure signal baseline model comprises the $f_{p13}$ component and $CP$, while the deformed model comprises all three components ($f_{p13}$, $f_{p22}$, and $f_{p32}$) and $CP$. From *Run 3* onwards, only the constraints of the $\varphi_{p3}$ parameter were modified (Table S2) while keeping the constraints of the rest of the parameters the same. This is to ensure the deformation produced by the $f_{p32}$ component is placed during or at the end of inspiration. The correct placement of the $f_{p32}$ component is accepted only when the difference between the end of the inspiration signal and the maximum of the $f_{p32}$ component response is positive- shown by the blue arrow. Otherwise, a red arrow indicates a negative difference. The estimated parameters for each Run are shown in Table S6.

**Table S6:** Estimated model parameters obtained from the optimization scheme for the breath representing Case 3 in Figure 3. In *Run 1*, only the baseline model (Equation 3 in the main text) parameters were estimated, while from *Run 2* onwards, all the model parameters were estimated.

| **Parameters** | **Case 3 - *Run 1*** | **Case 3 - *Run 2*** |
|---|---|---|
| $\theta$ | 0.2 | 0.2 |
| $\alpha_{v1}$ | 20.0 | 20.0 |
| $\beta_{v1}$ | 0.9 | 0.8 |
| $\varphi_{v1}$ | -0.8 | -0.8 |
| $\gamma_{v1}$ | 10.0 | 11.3 |
| $\gamma_{v2}$ | 5.0 | 20.0 |
| $A_{v1}$ | 477.9 | 446.9 |
| $\alpha_{v2}$ | 10.0 | 10.0 |
| $\beta_{v2}$ | 0.9 | 0.9 |
| $\varphi_{v2}$ | 2.0 | 2.0 |
| $A_{v2}$ | 200.0 | 200.0 |
| $\alpha_{p1}$ | 51.8 | 123.6 |
| $\beta_{p1}$ | 0.8 | 0.8 |
| $\varphi_{p1}$ | -0.7 | -0.7 |
| $\alpha_{p2}$ | - | 89.8 |
| $\beta_{p2}$ | - | 1.0 |
| $\varphi_{p2}$ | - | -1.2 |
| $\gamma_{p1}$ | 5.0 | 4.3 |
| $\gamma_{p2}$ | 3.4 | 3.4 |
| $A_{p1}$ | 5.9 | 3.5 |
| $A_{p2}$ | 0.0 | 1.7 |
| $CP$ | 8.2 | 8.2 |
| $\alpha_{p3}$ | - | 35.9 |
| $\beta_{p3}$ | - | 1.0 |
| $\varphi_{p3}$ | - | -0.3 |
| $A_{p3}$ | - | 5.2 |

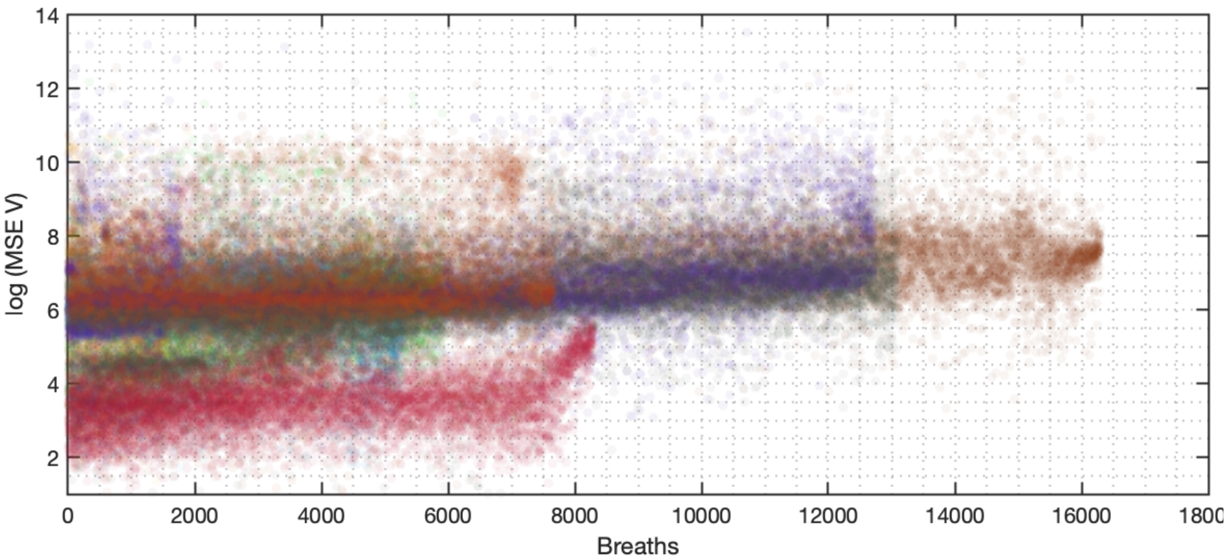


**Figure S10: Mean squares errors (MSE) produced by the VD-DLV volume model for all the estimated breaths.** The total number of estimated breaths shown here is 93,007. Each breath is represented by a dot. The mean value of log(MSE) and the corresponding standard deviation are 7.5 and 10.8, respectively.

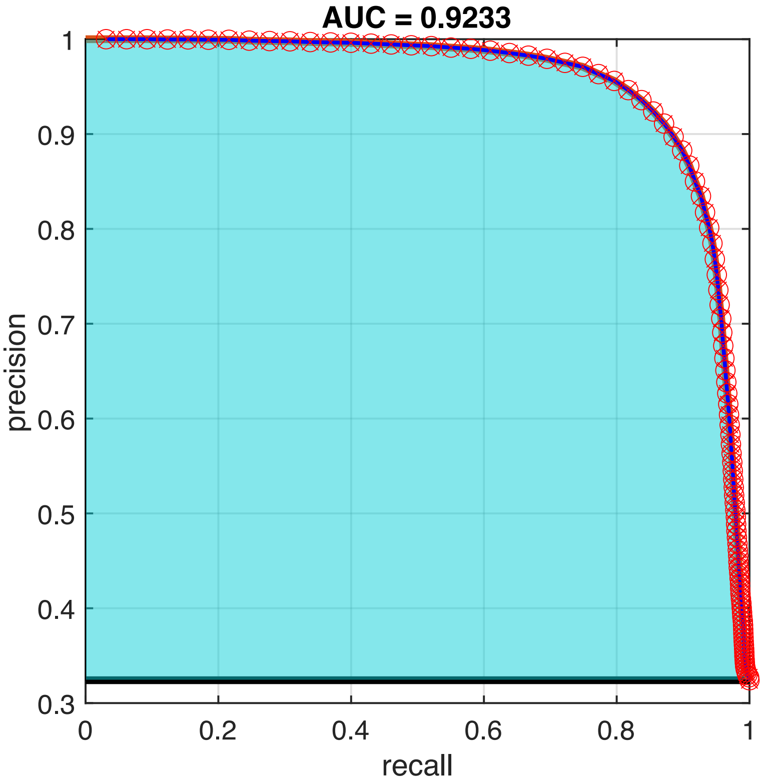


**Figure S11: Precision-recall curve.** Comparing the VD-DLV model classification of NL and FL-VD breaths with the human-guided ML algorithm (see Method). Area under the precision-recall curve is 0.92.

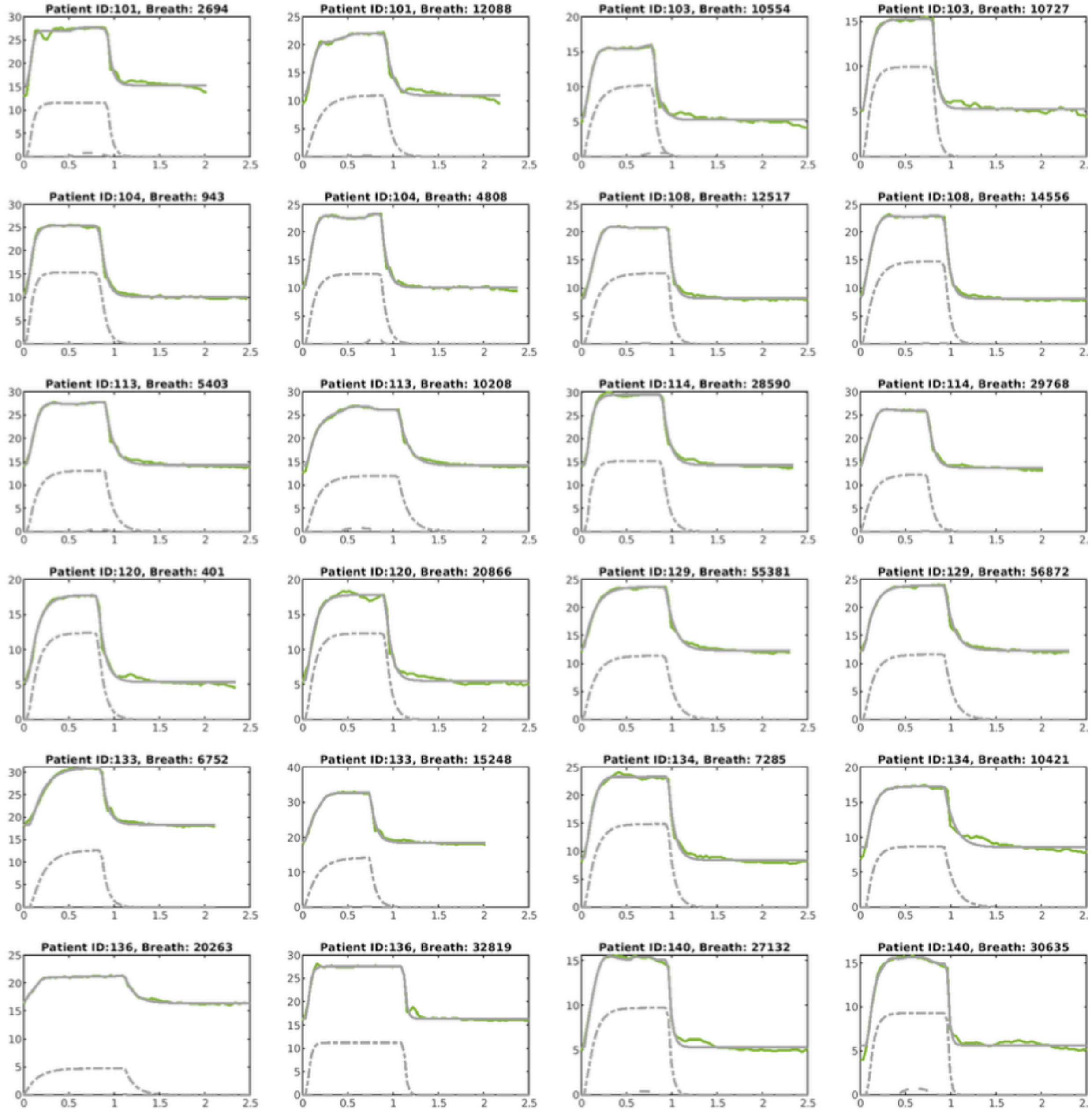


**Figure S12:** The pressure waveforms of some of the estimated normal breaths where the final model's response is shown in sold-grey lines, while the recorded data is shown in solid-colored lines. Response of the $f_{p13}$ and $f_{p32}$ components are shown in dashed-dashed lines and dashed-dot lines, respectively. Two breaths per patient are shown here, which were taken from the ten breaths that were randomly selected from each phenotype for each patient (see Fig. 5). Here, x and y axes represent time(sec) and pressure signals, respectively. The title of each subplot has the respective patient number ID and the breath number.

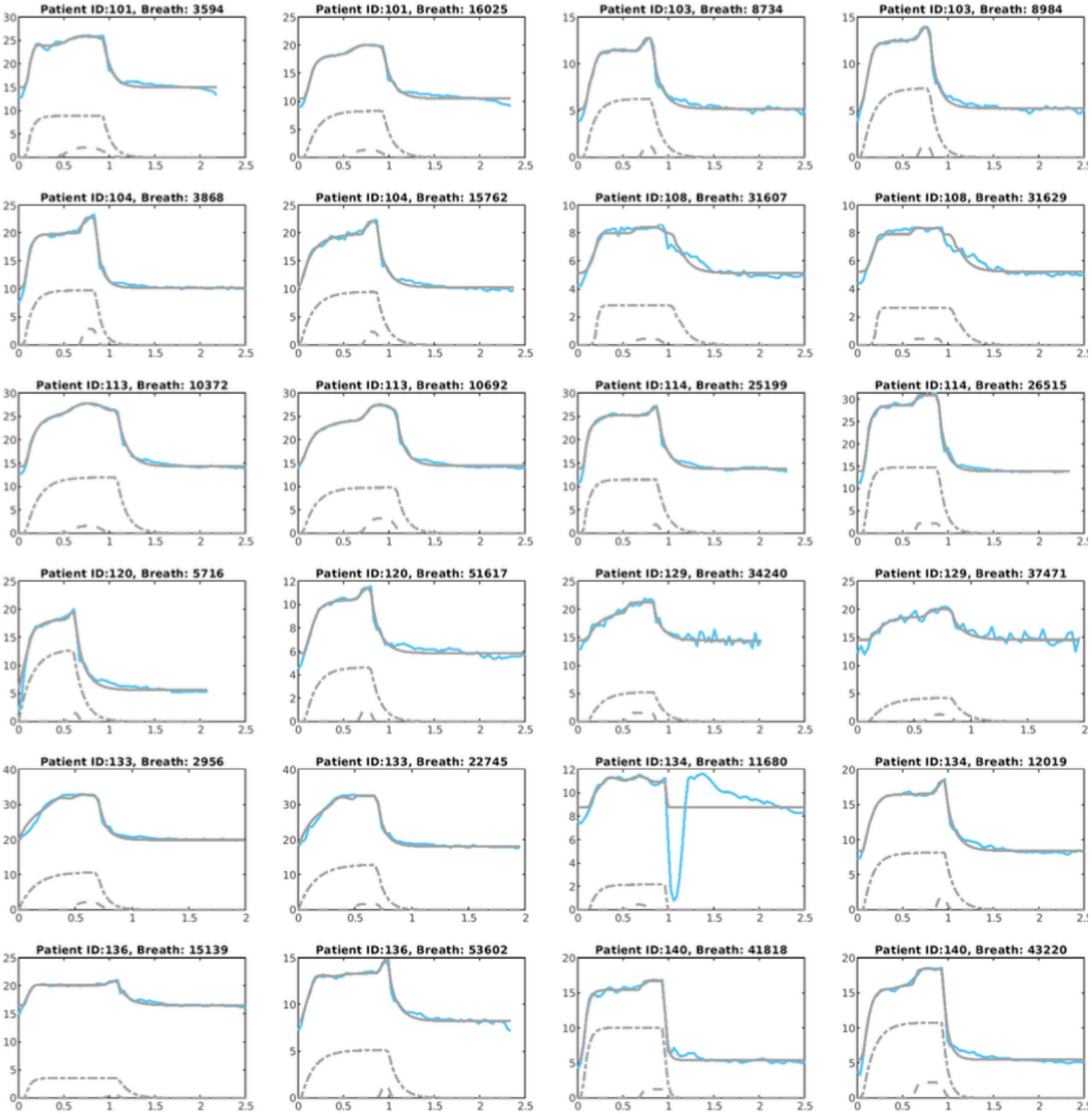


**Figure S13:** The pressure waveforms of some of the estimated mild flow limited breaths where the final model's response is shown in sold-grey lines, while the recorded data is shown in solid-colored lines. Response of the $f_{p13}$ and $f_{p32}$ components are shown in dashed-dashed lines and dashed-dot lines, respectively. Two breaths per patient are shown here, which were taken from the ten breaths that were randomly selected from each phenotype for each patient (see Fig. 5). Here, x and y axes represent time(sec) and pressure signals, respectively. The title of each subplot has the respective patient number ID and the breath number.

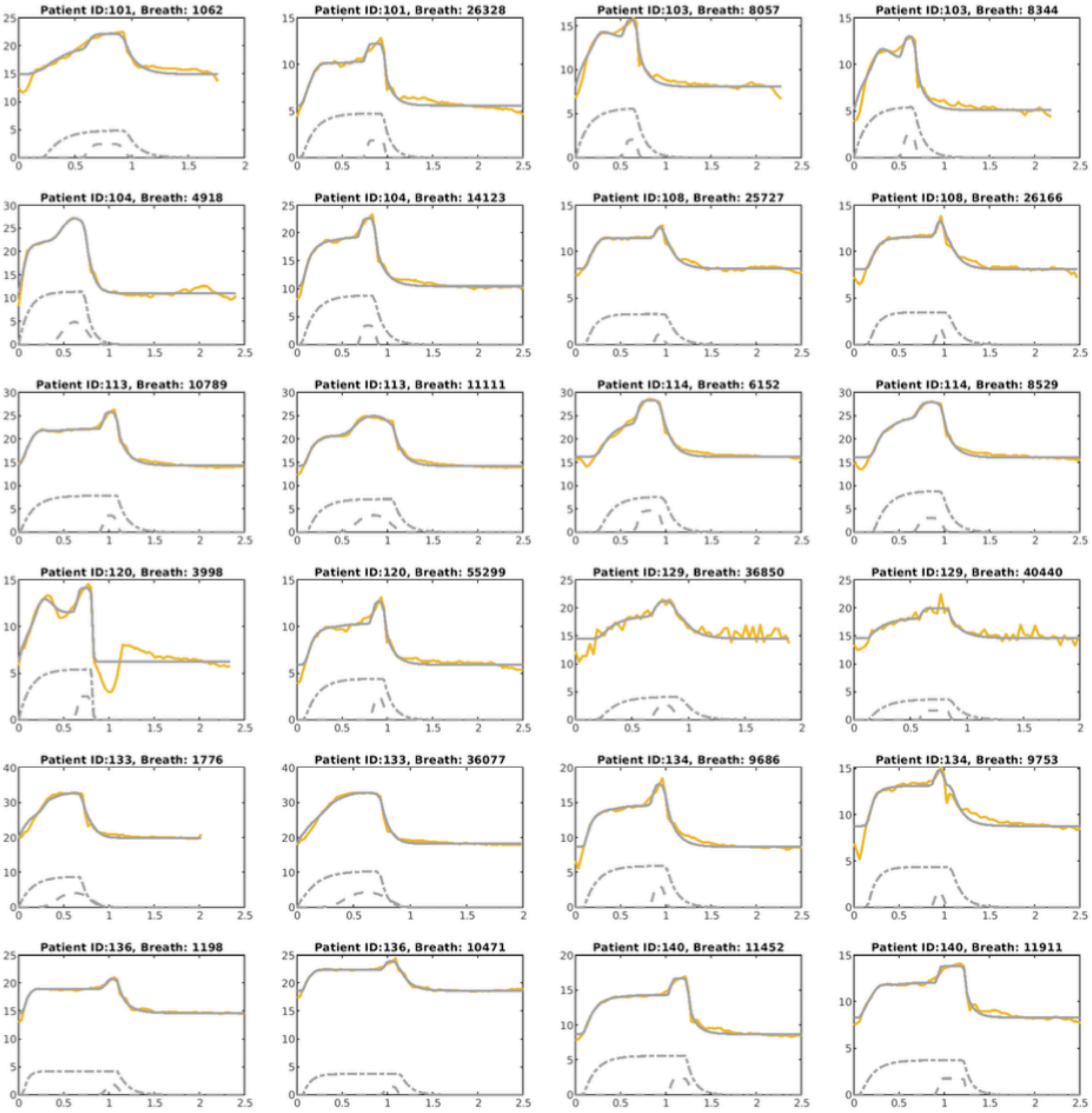


**Figure S14:** The pressure waveforms of some of the estimated intermediate flow limited breaths where the final model's response is shown in sold-grey lines, while the recorded data is shown in solid-colored lines. Response of the $f_{p13}$ and $f_{p32}$ components are shown in dashed-dashed lines and dashed-dot lines, respectively. Two breaths per patient are shown here, which were taken from the ten breaths that were randomly selected from each phenotype for each patient (see Fig. 5). Here, x and y axes represent time(sec) and pressure signals, respectively. The title of each subplot has the respective patient number ID and the breath number.

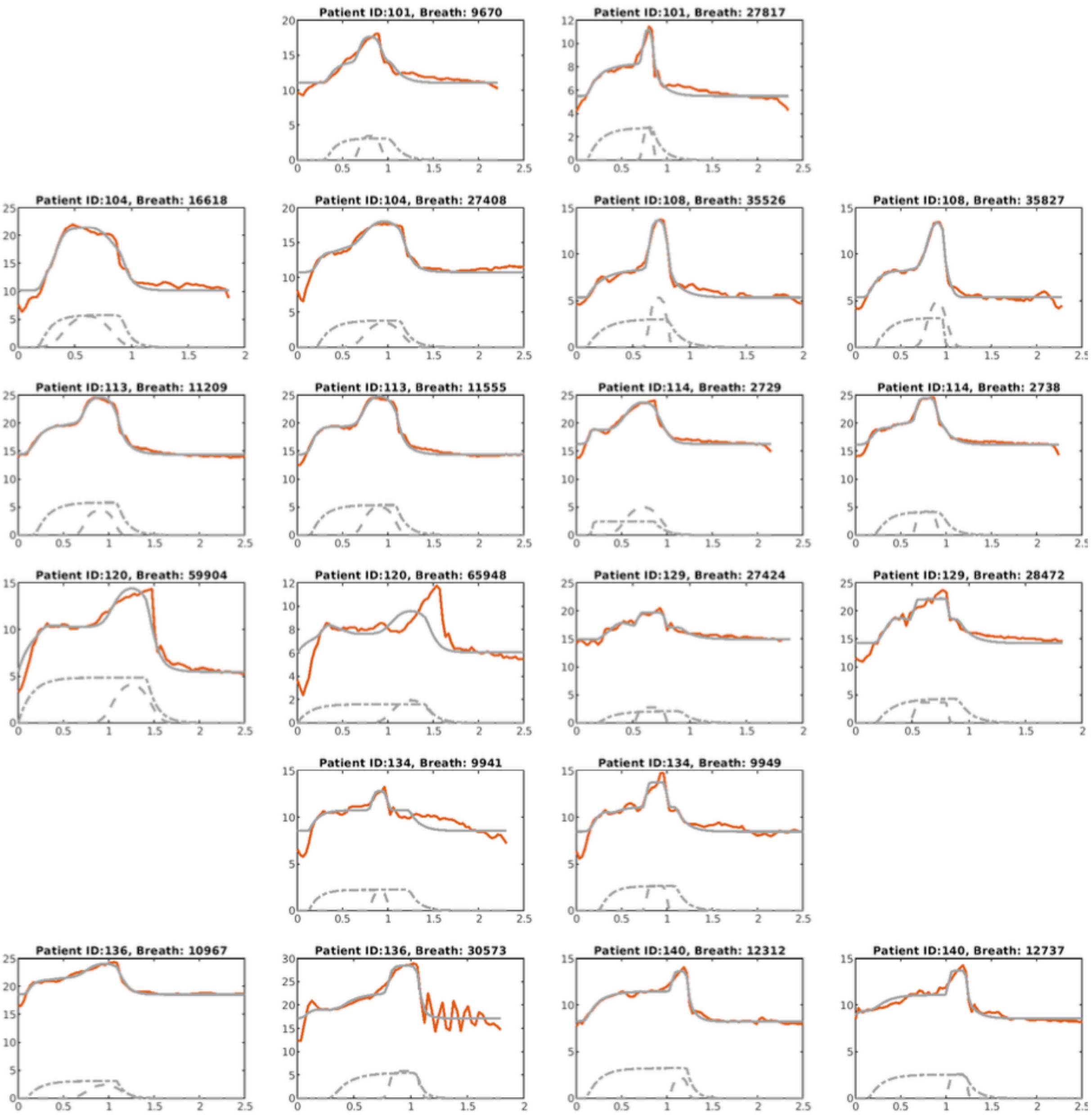


**Figure S15:** The pressure waveforms of some of the estimated severe flow limited breaths where the final model's response is shown in sold-grey lines, while the recorded data is shown in solid-colored lines. Response of the $f_{p13}$ and $f_{p32}$ components are shown in dashed-dashed lines and dashed-dot lines, respectively. Two breaths per patient are shown here, which were taken randomly from the manually reviewed selected breaths (see Fig. 5). Here, x and y axes represent time(sec) and pressure signals, respectively. The title of each subplot has the respective patient number ID and the breath number.
